# Does *in utero* HIV-exposure and the early nutritional environment influence infant development and immune outcomes? Findings from a pilot study in Pretoria, South Africa

**DOI:** 10.1101/19003889

**Authors:** Marina White, Ute D Feucht, Eleanor Duffley, Felicia Molokoane, Chrisna Durandt, Edana Cassol, Theresa Rossouw, Kristin L Connor

## Abstract

**Background:** As mother-to-child-transmission of HIV decreases, and the population of infants who are born HIV-exposed, but uninfected (HEU) continues to rise, there is growing need to understand the development and health outcomes of infants who are HEU to ensure that they have the healthiest start to life.

**Methods:** In a prospective cohort pilot study at Kalafong Hospital, Pretoria, South Africa, we aimed to determine if we could recruit new mothers living with HIV on antiretrovirals (ART; n=20) and not on ART (n=20), and new mothers without HIV (n=20) through our clinics to study the effects of HEU on growth, immune- and neuro-development in infants in early life, and test the hypothesis that infants who were HEU would have poorer health outcomes compared to infants who were HIV-unexposed, uninfected (HUU). We also undertook exploratory analyses to investigate relationships between the early nutritional environment, food insecurity, and infant development. Infant growth, neurodevelopment (Guide for Monitoring Child Development [GMCD]) and levels of monocyte subsets (CD14, CD16, and CCR2 expression [flow cytometry]) were measured in infants at birth and 12 weeks (range 8-16 weeks).

**Results:** We recruited 33 women living with HIV on ART, and 22 women living without HIV within four days of delivery from June-December 2016. 21 women living with HIV and 10 without HIV returned for a follow-up appointment at 12 weeks postpartum. The high mobility of this population presented major challenges to participant retention. Preliminary analyses revealed lower head circumference and elevated CCR2+ (% and median fluorescence intensity) on monocytes at birth among infants who were HEU compared to HUU. Maternal reports of food insecurity were associated with lower maternal nutrient intakes at 12 weeks postpartum and increased risk of stunting at birth for infants who were HEU, but not infants who were HUU.

**Conclusions:** Our small feasibility pilot study suggests that HEU may adversely affect infant development, and further, infants who are HEU may be even more vulnerable to the programming effects suboptimal nutrition *in utero* and postnatally. This pilot and preliminary analyses have been used to inform our research questions and protocol in our ongoing, full-scale study.

## Introduction

Maternal HIV infection profoundly affects maternal physiology and pregnancy outcomes. Annually, ∼1.3 million women living with HIV become pregnant^1,2^, and HIV infection in pregnancy is associated with increased risk of experiencing an adverse pregnancy outcome^3^, including preterm birth and maternal mortality^4,5^. Importantly, pregnancy and the postpartum period are two key times when HIV exposure may also have lasting impact on the fetus and infant, as fundamental structures, such as the brain, undergo rapid development *in utero*, and are thus vulnerable to infectious challenges such as HIV and drugs including antiretroviral therapies (ART)^6^.

Infants who are HIV-exposed and infected (HEI) show poorer motor, cognitive, language, and behavioural outcomes compared to controls as early as three months of age^7–9^. Importantly, global coverage of ART is increasing, reaching 80% of pregnant and breastfeeding women living with HIV in 2017^10^. This is true in South Africa, which faces the largest burden of HIV globally, and where an estimated 30% of pregnant women are living with HIV^11^. It is estimated that in 2018, 87% of women who were pregnant and living with HIV in South Africa had access to ART^12^ and the rate of mother-to-child HIV transmission (MTCT) was below 2% at birth^11^. Thus, as a result of declining MTCT, the number of infants being born who are HIV-exposed (*in utero* and during breastfeeding) but *uninfected* (HEU) is rising^13^.

Importantly, the extent to which HEU influences infant development is poorly understood, although evidence suggests that infants who are HEU have persistently altered motor and cognitive development^14–16^, albeit to a lesser extent than infants who are HEI. Further, in adults with HIV, markers of monocyte activation and altered frequencies of monocyte subsets are among some of the best predictors of non-AIDS associated co-morbid diseases^17,18^ and associate with increased neuro- and peripheral inflammation^19,20^. Infants who are perinatally-infected with HIV have increased monocyte activation between 4-15 weeks postpartum^21^ relative to infants who are HEU. However, it is less clear whether the distribution of monocyte subsets and their migratory potential are altered in infants who are HEU compared to infants who are HIV-unexposed, uninfected (HUU). Whether or not these alterations in HEU could explain some of the adverse neurodevelopmental and growth outcomes recorded in infants who are HEU is not well understood.

We conducted a prospective cohort pilot feasibility study in Pretoria, South Africa, to determine if we could recruit women through our clinics to study the effects of HEU on growth, immune- and neuro-development in infants in early life. Our first objectives were to test the feasibility of our research protocol and study design, and to and identify barriers to long-term follow up with mother-infant dyads. Our second objective was to perform exploratory analyses to test the hypothesis that infants who were HEU would have poorer growth and neurodevelopment, and alterations in monocyte subsets compared to infants who were HUU.

Lastly, as South Africa reports high rates of food and nutrition insecurity^22^, and the role of early life nutrition in infant health and development is well established^23–25^, it is also critical to understand how the early nutritional environment interacts with infectious exposures to influence developmental trajectories in infants who are HEU. Thus, our third objective was to explore the food security circumstances and dietary intakes of the study participants and relate these to our outcome measures. Together, objectives two and three were intended to inform the planning of our aims and analyses for a full-scale, future study.

## Methods

### Design, aims and setting

This pilot feasibility study was an observational, prospective clinical cohort study that took place at the obstetric unit at Kalafong Provincial Tertiary Hospital. We aimed to recruit new mothers living with HIV on antiretrovirals (ART; n=20) and not on ART (n=20), and new mothers without HIV (n=20) and their infants after delivery and follow them up in the early postpartum (PP) period, approximately 8-16 weeks after birth.

### Ethics

This study was approved by the Research Ethics Committee of the Faculty of Health Sciences of the University of Pretoria (185-2016) and the Carleton University Research Ethics Board (108870).

### Participant recruitment and eligibility

Eligible women were identified by a research nurse after delivery. Exclusion criteria included caesarean section delivery, pregnancy complications (including gestational diabetes mellitus, multiple gestations), or antibiotic exposure during labour or delivery and/or the postpartum period. Women were also ineligible to participate if they were from other regions and would find it difficult to come back for follow-up. All infants exposed to HIV were tested for infection at birth and 12 weeks postpartum. As we were interested in exploring the effects of HIV-exposure without infection on infant growth, neurodevelopment, and immune outcomes, if an infant was determined to have HIV, the mother-infant dyad was subsequently excluded from the study analyses.

### Data collection

#### Maternal pregnancy and postnatal environment data

After delivery, a retrospective medical chart review was conducted to extract antenatal data. This included maternal characteristics (age at conception, parity, gravidity, smoking status, weight during pregnancy); medication use during pregnancy (including antibiotic exposure); illness/infections during pregnancy; and pregnancy outcomes (gestation length). At the postpartum follow up visit, mothers completed a questionnaire to assess breastfeeding practices, maternal lifestyle factors (including alcohol and smoking), and nutrition (including vitamin supplements, food security, and a 24-hour dietary recall). If any visits to clinics or hospitals occurred between birth and the follow up visit, the patient-retained child health record (Road to Health Booklet^26^) of the infant was examined to extract data on infant weight, history of illness and medication use.

#### Infant outcomes at birth and 12 weeks postpartum

Infant weight, length, and abdominal and head circumference were measured at birth and 12 weeks postpartum. Apgar score at one and five minutes was obtained. Infant anthropometry were age- and sex-standardised using World Health Organization (WHO) growth standards (WHO Anthro software [v 3.2.2, January 2011])^27^. A brain weight estimate was calculated using an equation derived by the National Institute of Neurological and Communicative Disorders and Stroke’s Collaborative Perinatal Project^28^: *brain weight (g) = 0*.*037 × head circumference (cm)*^*2*.*57*^. The brain weight estimate was used to calculate the infant brain-to-body weight ratio (BBR)^29^: *BBR = 100 × (brain weight estimate [g])/(birth weight [g])*. Weight gain from birth to 12 weeks postpartum (kg/day) was calculated using the weight of an infant at birth and follow up, and the days alive since birth at follow up: *(weight at 12 weeks postpartum [kg] – weight at birth [kg])/number of days alive*.

#### Infant monocyte subsets

All infants who were HEU underwent a blood draw at birth and again at 12 weeks for HIV testing. Blood from this routine draw was obtained within 4 days of birth and again at 12 weeks and used to quantify the surface markers CD14, CD16 and CCR2 for monocyte subset identification^30^. Using the Gallios flow cytometer (3 laser, 10 colour configuration; Beckman Coulter, Miami, FL, USA), CD14 expression on PBMCs and CD16 and CCR2 expression (% and median fluorescence intensity [MFI]) on monocyte subsets was evaluated within four days of birth and at 12 weeks of age. The Kaluza V1.0 Acquisition software (Beckman Coulter, Miami, FL, USA) was used for data acquisition and post-acquisition data analysis was performed using Kaluza Analysis software (Version V3.1; Beckman Coulter, Miami, FL, USA). Flow-Check Pro fluorosphere were acquired daily prior to sample analysis to ensure optimal laser alignment and instrument performance. Single colour staining tubes (i.e. sample stained with individual monoclonal antibodies) were used to setup the protocol and calculate the colour compensation values. After setup and protocol/template verification, the instrument settings (voltages, gains, threshold and colour compensation settings) were kept the same throughout the study. The reagent list, including lasers and detectors used, and compensation matrix are presented in Supplementary tables S1 and S2, and the analysis approach and gating strategy are described in Supplementary figures S1 and S2. Classical (CD14^++^CD16^−^), intermediate (CD14^++^CD16^+^) and non-classical (CD14^+^CD16^+^) monocyte subsets were identified^31^.

#### Infant neurodevelopment

The Guide for Monitoring Child Development (GMCD)^32^ assesses expressive and receptive language, play activities, relating and response behaviour, and fine and large movement. The GMCD was developed for use in low- and middle-income countries to assess infants from 1 to 24 months postpartum, and involves the researcher asking the child’s caregiver a series of open-ended questions relating to the child’s development. An assessment for each infant was carried out once between 8-16 weeks postpartum. Infants who were 1-3 months of age (1 month to 2 months and 30 days) were assessed on milestones listed in the 1-3 month category, and infants who were 3-5 months (3 months+1 day to 4 months+30 days) were assessed for milestones listed in both the 1-3 and 3-5 month columns. Infants who were premature (<37 weeks) were age-corrected to term. The GMCD has been standardised and validated for international use in a sample of approximately 12,000 children from 4 diverse countries, namely South Africa, Argentina, India and Turkey^33^. The proportion of infants having attained all milestones (compared to not having attained all milestones)^34^ in their age category (1-3 months, or 3-5 months) was quantified.

#### Maternal reports of food security, dietary recall, and infant feeding patterns

A questionnaire was developed to collect maternal reports of food security. Mothers were asked if, in the past 12 months, the following were ‘often true’, ‘sometimes true’, or ‘never true’: 1. They and other household members worried that food would run out before they got money to buy more, 2. the food that they and other household members bought just didn’t last, and there wasn’t any money to get more, and 3. they and other household members couldn’t afford to eat balanced meals. For the purpose of exploratory analyses and due to the small sample size of the pilot, maternal reports of ‘often true’ and ‘sometimes true’ were grouped together for analyses as ‘experiences food insecurity’ and compared with ‘never true’ responses.

Maternal dietary recall data collected a detailed account of all food and drink consumed in the day prior to the follow up appointment. Dietary recall data were analysed using FoodFinder3^35^, a dietary analysis software programme developed by the South African Medical Research Council, specific to the nutrient composition of foods in South Africa. The estimated average requirements (EARs) and tolerable upper levels (TULs) for available nutrients from the Institute of Medicine Dietary Reference Intakes were used to evaluate the nutritional adequacy of reported maternal diets^36^. These reference intakes have been used previously to evaluate diet composition in various South African cohorts^37^. A dietary diversity score (DDS) was calculated as an additional measure of diet quality using nine food groups (1. Cereals, roots and tubers, 2. Vegetables and fruits rich in Vitamin A, 3. Other fruit, 4. Other vegetables, 5. Legumes, 6. Meat, poultry and fish, 7. Dairy, 8. Eggs, and 9. Fats and oils) as previously validated and described in South African cohorts^38,39^. Each food group was only counted once.

At follow up, mothers reported whether they were, or had ever, exclusively breastfed their infants. If the infants were currently receiving formula, the mothers provided the age at which formula had been introduced.

### Statistical analyses

Data were analysed using JMP 14.0. One-way analysis of variance (ANOVA, Kruskal-Wallis/Wilcoxon test for non-parametric data, or Welch’s test normal data with unequal variance) and adjusted multiple regression models were used to explore: 1. infant anthropometry at birth and 12 weeks postpartum, 2. Apgar scores (one and five minutes), 3. levels of total monocytes, monocyte subsets and CCR2 expression assessed within 4 days of birth and at 12 weeks postpartum, and 4. number of GMCD milestones attained for infants who were HEU compared to infants who were HUU. We also explored maternal dietary intake nutrient levels for mothers living with and without HIV, and possible relationships between household food insecurity and infant outcomes through comparisons of infant outcomes at birth and 12 weeks for infants whose mothers reported on food security.

Using Fisher’s exact test (2-tail), we compared the probability of: 1. attaining all age-appropriate GMCD milestones, and 2. stunting at birth or 12 weeks of age for infants who were HEU or HUU. We also explored relationships between maternal reports of food insecurity and probability of 1. stunting at birth or 12 weeks of age, 2. attaining all age-appropriate GMCD milestones, and 3. exclusively breastfeeding at follow up.

Maternal age and weight at delivery, and infant gestational age, sex, and age (days) at their follow up appointment were included as covariables in adjusted analysis. Outliers were excluded where measurement error was clear, including for one participant for %CCR2-positive classical monocytes at birth, and one for %CCR2-positive non-classical monocytes at 12 weeks. Data for infant outcomes below are presented as unadjusted means (SD) or medians (IQR) with p value from ANCOVA (*p*<0.05).

To determine whether or not the study cohort at follow up was representative of the cohort at birth, one-way analysis of variance compared all outcomes at birth for infants who were, versus were not, present at follow up, within the groups of infants who were HEU and HUU. Data below that compares outcomes in infants who were lost to follow up to those who were not are presented as unadjusted means (SD) or medians (IQR) and p values (*p*<0.05).

## Results

### Recruitment and participant study groups

Study recruitment took place between June and December 2016. By March 2017, all follow up data had been collected. An overview of the pilot study design, methods, and participation is described in Fig 1. At the end of recruitment, we recruited 55 women within four days of delivery: 33 living with HIV on ART and 22 living without HIV. Due to changed treatment policies, all women living with HIV were already on ART when they enrolled into the study. Secondly, the time set out for recruitment lapsed before we could enrol the planned numbers of women living without HIV. One infant whose mother was living with HIV tested positive for HIV at birth and again at 12 weeks. This mother-infant dyad was subsequently excluded from the study analyses, making the final groups at delivery: HIV-uninfected, n=22; HIV-infected, n=32. Attrition at follow up was high, with 31.25% of women living with HIV and 54.5% of those living without HIV not attending a follow up appointment. Thus, 21 women living with HIV and 10 without were followed-up at one timepoint 12 weeks (range 8-16 weeks) postpartum. Multiple attempts were made to contact women who missed follow up appointments via the phone numbers provided at recruitment. The high mobility of this population presented major challenges to participant retention.

**Figure 1.**
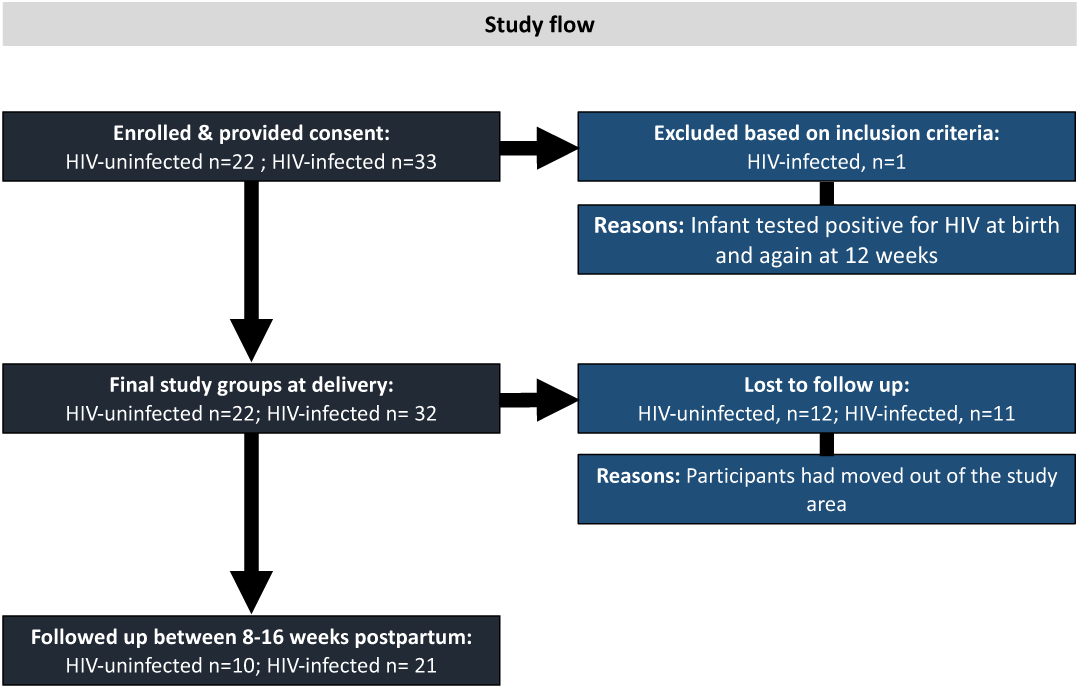
Study flow diagram. 33 women living with HIV on ART, and 22 women living without HIV were recruited within four days of delivery from June-December 2016. One infant whose mother was living with HIV tested positive for HIV at birth and again at 12 weeks and was excluded from the study analyses, making the final groups at delivery: HIV-uninfected, n=22; HIV-infected, n=32. 31.25% of women living with HIV and 54.5% of those living without HIV did not attend a follow up appointment. 21 women living with HIV and 10 without were followed-up at one timepoint 12 weeks (range 8-16 weeks) postpartum. The high mobility of this population presented major challenges to participant retention.

### Maternal cohort characteristics

Maternal cohort characteristics are described in Table 1. There were no differences in maternal age, gravidity, parity, level of education or weight at delivery between mothers living with vs. without HIV. All mothers identified as non-cigarette smokers, however, one woman reported consuming snuff.

**Table 1.** Maternal cohort characteristics.

|  | HIV uninfected<br>(n=22) | HIV infected<br>(n=32) | p<br>value |
| --- | --- | --- | --- |
| **Maternal characteristics** |  |  |  |
| Weight at delivery (kg) | 60.1 (55.0, 80.3) | 60.4 (56.0, 69.0) | NS |
| Age (years) | 28.1 ± 7.31 | 30.8 ± 5.91 | NS |
| Gravidity (n) | 2.00 (1.50, 4.00) | 3.00 (2.00, 3.00) | NS |
| Parity (n) | 2.00 (1.00, 3.00) | 2.00 (1.00, 3.00) | NS |
| Level of education (n) |  |  |  |
| Primary | 1 | 6 |  |
| Secondary | 17 | 24 |  |
| Post-secondary | 4 | 1 |  |
| Not indicated | - | 1 |  |
| Current ART (n) |  |  |  |
| TDF, FTC, EFV | - | 28 |  |
| AZT, 3TC, LPV/r | - | 1 |  |
| None |  | 3 |  |

Data are means ± SD (ANOVA; normal distribution/equal variance) or median (IQR; Kruskal-Wallis/Wilcoxon test for non-parametric data, or Welch's test for normal data/unequal variance). NS = non-significant; TDF = Tenofovir; FTC = emtricitabine; EFV = efavirenz; AZT = azidothymidine; 3TC = lamivudine; LPV/r = ritonavir-boosted lopinavir.

### Infant outcomes at birth

#### Infant birth outcomes and demographics

Infant cohort characteristics are presented in Table 2. Among infants who were HUU, 45.5% were male, while 34.4% of infants who were HEU were male. Preterm birth (<37 weeks) occurred in 5/54 (9.26%) pregnancies. Two preterm infants were HUU, born at 36 and 35 weeks, and three were HEU, with two born at 36 weeks, and one at 35 weeks.

**Table 2.** Infant cohort characteristics and anthropometric, immune, and developmental outcomes at birth and 8-16 weeks postpartum.

|  | HIV-unexposed-<br>uninfected<br>n=22 | HIV-exposed-<br>uninfected<br>n=32 | p value<br>(unadjusted) | p value<br>(adjusted) |
| --- | --- | --- | --- | --- |
| **Infant outcomes at birth** |  |  |  |  |
| Infant sex (% male) | 45.5 | 34.4 | NS | - |
| Gestational age at delivery (weeks) | 38.4 ± 1.56 | 38.7 ± 1.69 | NS | - |
| <37 weeks GA (n) | 2 | 3 | NS | - |
| <37 weeks GA (weeks) | 35.5 (35.0, 36.0) | 36.0 (35.0, 36.0) | NS | - |
| **Infant anthropometry<sup>1</sup>** |  |  |  |  |
| Weight z-score | -0.57 ± 1.15 | -0.76 ± 0.96 | NS | NS |
| Length z-score | -1.33 (-3.20, 0.45) | -1.65 (-2.33, -0.94) | NS | NS |
| BMI z-score | -0.15 (-1.76, 1.80) | 0.09 (-0.81, 0.97) | NS | NS |
| Head circumference z-score | -0.33 ± 1.19 | -1.58 ± 1.22 | <0.001 | <0.001 |
| Brain-to-body weight ratio | 10.2 (9.77, 11.3) | 9.60 (8.88, 10.4) | NS | NS |
| Stunting (n [%]) | 7 (33.3) | 14 (43.8) | NS | - |
| **Apgar score** |  |  |  |  |
| 1 minute | 9.00 (8.00, 9.00) | 9.00 (8.00, 9.00) | NS | NS |
| 5 minute | 9.00 (9.00, 10.0) | 9.00 (9.00, 9.00) | NS | NS |
| **Immune measures** |  |  |  |  |
| Total CD14+ | n=21<br>10.5 (8.19, 12.2) | n=32<br>8.76 (6.23, 12.0) | NS | NS |
| %CCR2-positive CD14+ monocytes | 82.9 ± 6.91 | 86.6 ± 4.85 | 0.03 | NS |
| **Monocyte sub-populations (%)** |  |  |  |  |
| Classical (CD14++/CD16-) | 67.8 ± 13.7 | 70.8 ± 13.3 | NS | NS |
| Intermediate (CD14++/CD16+) | 14.7 (9.83, 21.2) | 12.9 (9.35, 17.0) | NS | NS |
| Non-classical (CD14+/CD16+) | 8.40 (4.26, 15.4) | 8.44 (4.17, 13.0) | NS | NS |
| **%CCR2-positive monocyte sub-populations** |  |  |  |  |
| Classical (CD14++/CD16-) | 94.7 (92.2, 97.2) | 97.4 (95.2, 98.4) | 0.01 | 0.01 |
| Intermediate (CD14++/CD16+) | 60.4 (51.4, 71.9) | 68.2 (62.3, 80.9) | NS | NS |
| Non-classical (CD14+/CD16+) | 8.05 (4.00, 15.4) | 6.88 (3.55, 15.1) | NS | NS |
| **CCR2 MFI on monocyte sub-populations** |  |  |  |  |
| Classical (CD14++/CD16-) | 5.25 (4.49, 6.63) | 6.29 (5.59, 6.81) | 0.04 | NS |
| Intermediate (CD14++/CD16+) | 2.33 (2.16, 2.65) | 2.87 (2.34, 3.48) | 0.003 | 0.04 |
| Non-classical (CD14+/CD16+) | 1.65 (1.55, 1.92) | 1.70 (1.49, 2.22) | NS | NS |
| **Infant outcomes at 12 weeks postpartum** |  |  |  |  |
| Infant sex (% male) | n=10<br>50.0 | n=21<br>33.3 | NS | - |
| Infant age at follow up (weeks) | 10.4 (10.1, 12.1) | 12.0 (10.1, 13.3) | NS | - |
| EBF at follow up (n [%]) | 4 (40.0) | 14 (81.0) | NS | - |
| **Infant anthropometry<sup>a</sup>** |  |  |  |  |
| Head circumference z-score | -0.73 (1.40, -0.58) | -1.47 (-2.17, -0.97) | 0.03 | NS |
| Weight z-score | 0.12 (-1.02, 0.56) | -0.46 (-0.93, 0.02) | NS | NS |
| Length z-score | -0.95 (-2.74, -0.27) | -1.18 (-1.69, -0.62) | NS | NS |
| BMI z-score | 0.44 ± 1.92 | 0.19 ± 1.12 | NS | NS |
| Brain-to-body weight ratio | 7.39 (6.67, 8.04) | 7.59 (7.08, 7.83) | NS | NS |
| Weight gain (birth to 12 weeks PP; kg/day) | 0.04 ± 0.01 | 0.03 ± 0.01 | NS | NS |
| Stunting at 12 weeks postpartum<sup>1</sup> (n [%]) | 3 (30.0) | 2 (9.52) | NS | - |
| **Immune measures** |  |  |  |  |
| Total CD14+ | n=10<br>7.40 (5.69, 9.78) | n=17<br>7.16 (5.69, 8.98) | NS | NS |
| %CCR2-positive CD14+ monocytes | 80.3 (74.9, 85.3) | 76.0 (63.5, 80.2) | NS | NS |
| **Monocyte sub-populations (%)** |  |  |  |  |
| Classical (CD14<sup>++</sup>/CD16<sup>-</sup>) | 68.0 (67.5, 75.3) | 64.1 (57.3, 72.9) | NS | NS |
| Intermediate (CD14<sup>++</sup>/CD16<sup>+</sup>) | 8.49 (6.61, 13.7) | 12.6 (8.95, 22.3) | NS | NS |
| Non-classical (CD14<sup>+</sup>/CD16<sup>+</sup>) | 16.7 ± 4.94 | 19.4 ± 5.85 | NS | NS |
| %CCR2-positive monocyte sub-populations |  |  |  |  |
| Classical (CD14<sup>++</sup>/CD16<sup>-</sup>) | 93.3 (89.3, 96.6) | 94.7 (93.5, 96.2) | NS | NS |
| Intermediate (CD14<sup>++</sup>/CD16<sup>+</sup>) | 44.8 ± 23.1 | 56.2 ± 12.2 | NS | NS |
| Non-classical (CD14<sup>+</sup>/CD16<sup>+</sup>) | 1.59 (0.82, 3.09) | 3.42 (1.70, 4.32) | NS | NS |
| CCR2 MFI on monocyte sub-populations |  |  |  |  |
| Classical (CD14<sup>++</sup>/CD16<sup>-</sup>) | 4.67 (3.83, 6.04) | 4.46 (4.03, 4.89) | NS | NS |
| Intermediate (CD14<sup>++</sup>/CD16<sup>+</sup>) | 2.19 (1.95, 3.14) | 2.22 (1.89, 2.49) | NS | 0.04 |
| Non-classical (CD14<sup>+</sup>/CD16<sup>+</sup>) | 1.55 (1.40, 2.52) | 1.64 (1.40, 1.93) | NS | NS |

Data are presented means ± SD (ANOVA) or median (IQR; Kruskal-Wallis/Wilcoxon test for non-parametric data, or Welch's test for normal data/unequal variance) with p values from one-way analysis of variance and adjusted multiple variable regression models. Differences in proportions for infant sex, <37 weeks gestational age, % EBF (exclusively breastfed) at follow up, and risk of stunting for HUU vs. HEU infants were assessed by Fisher's exact test (2-tail). <sup>1</sup>All infant anthropometric measures are standardised according to WHO child growth standards<sup>29</sup>. <sup>2</sup>Stunting is determined by <-2 SD length-for-age standardised z-score according to WHO child growth standards (WHO | WHO Anthro (version 3.2.2, January 2011) and macros, 2017). PP = postpartum.
<sup>a</sup>Infant anthropometry was adjusted for maternal age, maternal weight, gestational age
<sup>b</sup>BBR, Apgar score (one and five minutes), immune measures at birth, and weight gain (birth to 12 weeks PP; kg/day) were adjusted for maternal age, maternal weight, gestational age, infant sex

#### Anthropometric measures and Apgar scores

Exploratory analyses revealed lower head circumference-for-age z-scores (-1.58 ± 1.22 vs. -0.33 ± 1.19, p<0.001; Fig 2D, Table 2) at birth among infants who were HEU compared to HUU. There were no differences between infants who were HEU and HUU for BMI, length-for-age and weight-for-age z-scores, BBR at birth, or Apgar scores (one minute, five minutes) (Fig 2, Table 2).

**Figure 2.**
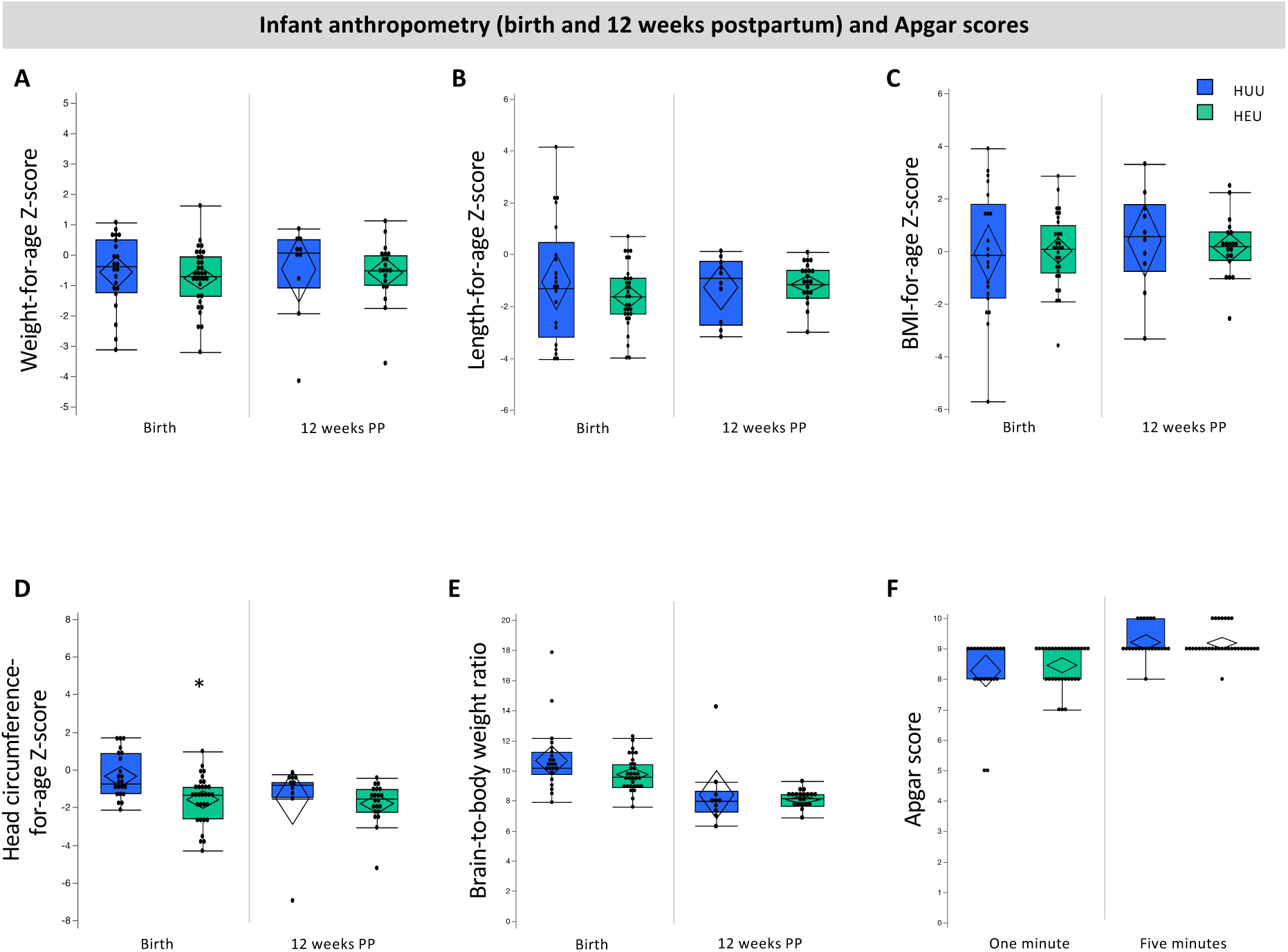
Relationships between in utero HIV-exposure and infant anthropometry at birth and 12 weeks (A-E) and Apgar scores (F). Infants who were HEU (n=32) had lower head circumference Z-scores at birth (D; p<0.001) compared to infants who were HUU (n=22). There were no other differences between infants who were HEU compared to HUU for anthropometry at birth or 12 weeks of age. Outlier box plots are measured anthropometry and Apgar scores (quartiles, median lines and 95% confidence diamonds, *p<0.05 [ANCOVA]). HUU = HIV-unexposed, uninfected; HEU = HIV-exposed, uninfected; CI = confidence interval.

#### Immune measures

There were no differences between infants who were HEU vs. HUU in the relative frequency of total monocytes (% CD14+ PBMC), CCR2 expression by total CD14+ monocytes or the monocyte subsets (Table 2). Infants who were HEU had elevated %CCR2-positive classical monocytes (97.4 [95.2, 98.4] vs. 94.7 [92.2, 97.2], p=0.01; Fig 3A, Table 2) at birth. No differences were observed for %CCR2-positive intermediate - or non-classical monocytes between the two groups at birth (Fig 3). Median fluorescence intensity (MFI) is indicative of the level of cell surface expression and a higher CCR2 MFI were observed for intermediate (2.87 [2.34, 3.48] vs. 2.33 [2.16, 2.65], p=0.04; Fig 3E) at birth in infants who were HEU when compared to infants who were HUU. No significant differences in CCR2 expression levels were observed in classical or non-classical monocytes at birth in infants who were HEU compared to HUU (Fig 2F, Table 2).

**Figure 3.**
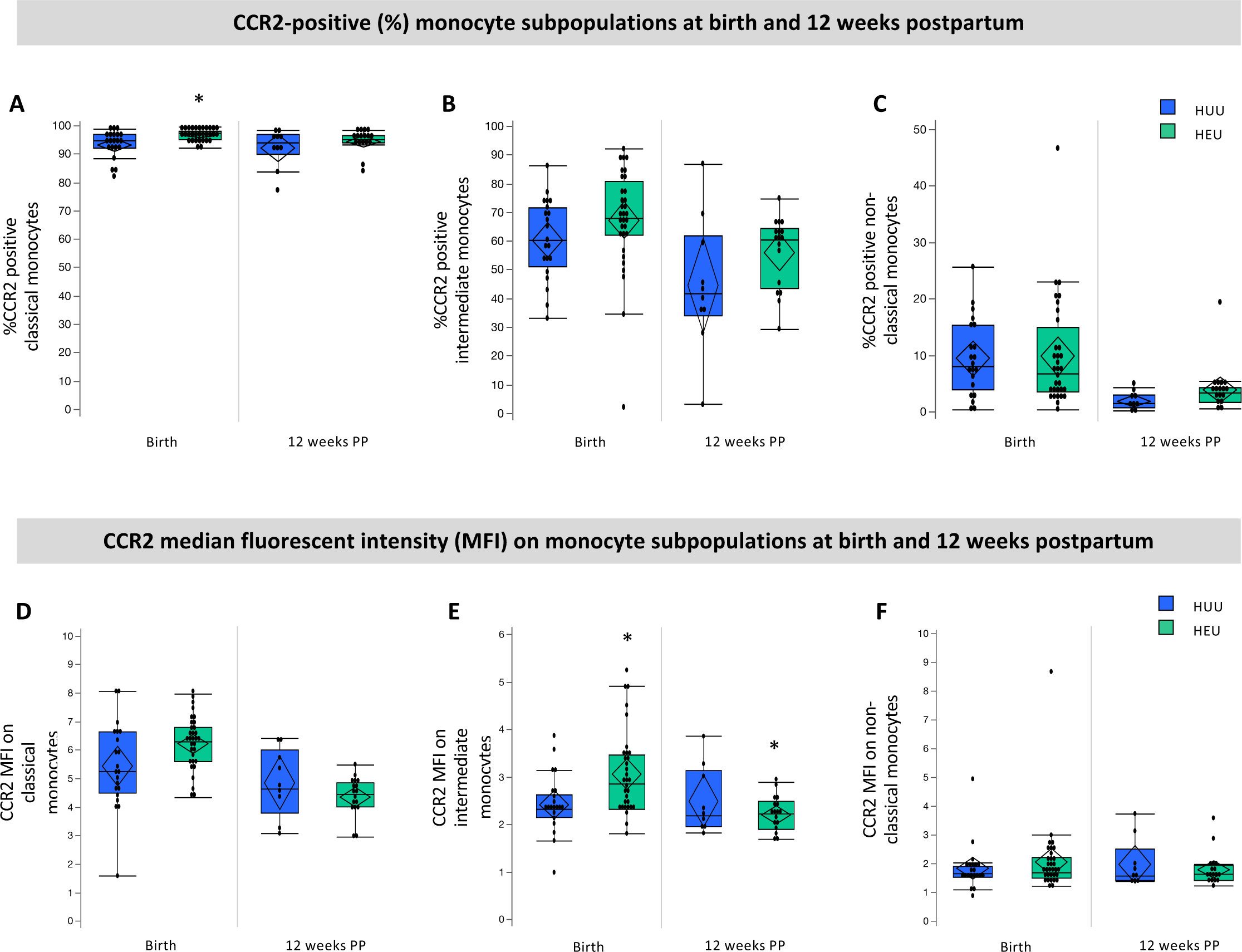
CCR2 expression by monocyte subpopulations within four days of birth and at 12 (±4) weeks of age. Infants who were HEU (n=32) had elevated %CCR2-positive classical monocytes (A; p=0.01), and CCR2 expression (MFI) on intermediate monocytes at birth (E; p=0.03) and 12 weeks (E; 0.04) compared to infants who were HUU (n=21). Data are measured as proportion of monocyte sub-populations expressing CCR2 (%) and the average (median) levels of expression per cell (MFI) on monocyte subpopulations. Outlier box plots are quartiles, median lines and 95% confidence diamonds, *p<0.05 [ANCOVA]). HUU = HIV-unexposed, uninfected; HEU = HIV-exposed, uninfected; MFI = median fluorescent intensity (MFI); CI = confidence interval.

### Infant outcomes at 12 weeks postpartum

#### Demographics

At follow up, 50.0% of infants who were HUU were male, while 33.3% of infants who were HEU were male (Table 2). There were no differences between infants who were HUU and HEU for age at follow up.

#### Anthropometric measures

There were no differences between infants who were HEU and HUU for age- and sex-standardised anthropometric measures at follow up (Fig 2, Table 2), and no differences in weight gain from birth to 12 weeks postpartum (Table 2).

#### Immune measures

Infants who were HEU had elevated CCR2 MFI on intermediate monocytes compared to HUU at 12 weeks postpartum (2.22 [1.89, 2.49] vs. 2.19 [1.95, 3.14], p=0.04; Fig 3E), but not on classical or non-classical monocytes. There were no differences between infants who were HEU compared to HUU at 12 weeks postpartum for %CD14^+^ PMBCs, mean monocyte subsets or %CCR2 on all monocyte subsets (Table 2).

#### GMCD

The percentage of infants in each group (HUU compared to HEU) who had achieved all age-appropriate milestones at 1-3 or 3-5 months postpartum are presented (Fig 4). One infant who was HUU did not undergo a GMCD developmental assessment at follow up. 23/31 (74.19%) of infants aged 8-16 weeks had a 1-3 month GMCD assessment. Of these infants, 15/23 (65.2%) were HEU. At 1-3 months, infants who were HUU had attained age-appropriate milestones for receptive language, large movement and play activity milestones in lower proportions (-10.0%, -10.0%, and -22.5%, respectively) than the GMCD international standardised sample (Fig 4A). Infants who were HEU who were 1-3 months of age had attained all age-appropriate milestones for 1-3 months postpartum. We found no association between HIV-exposure group and probability of attaining all age-appropriate milestones (compared to not attaining) in any GMCD neurodevelopmental theme at 1-3 months (Fisher’s exact). All infants who were 3-5 months at follow up had attained all 1-3 month milestones for all themes, consistent with the GMCD international standardised sample.

**Figure 4.**
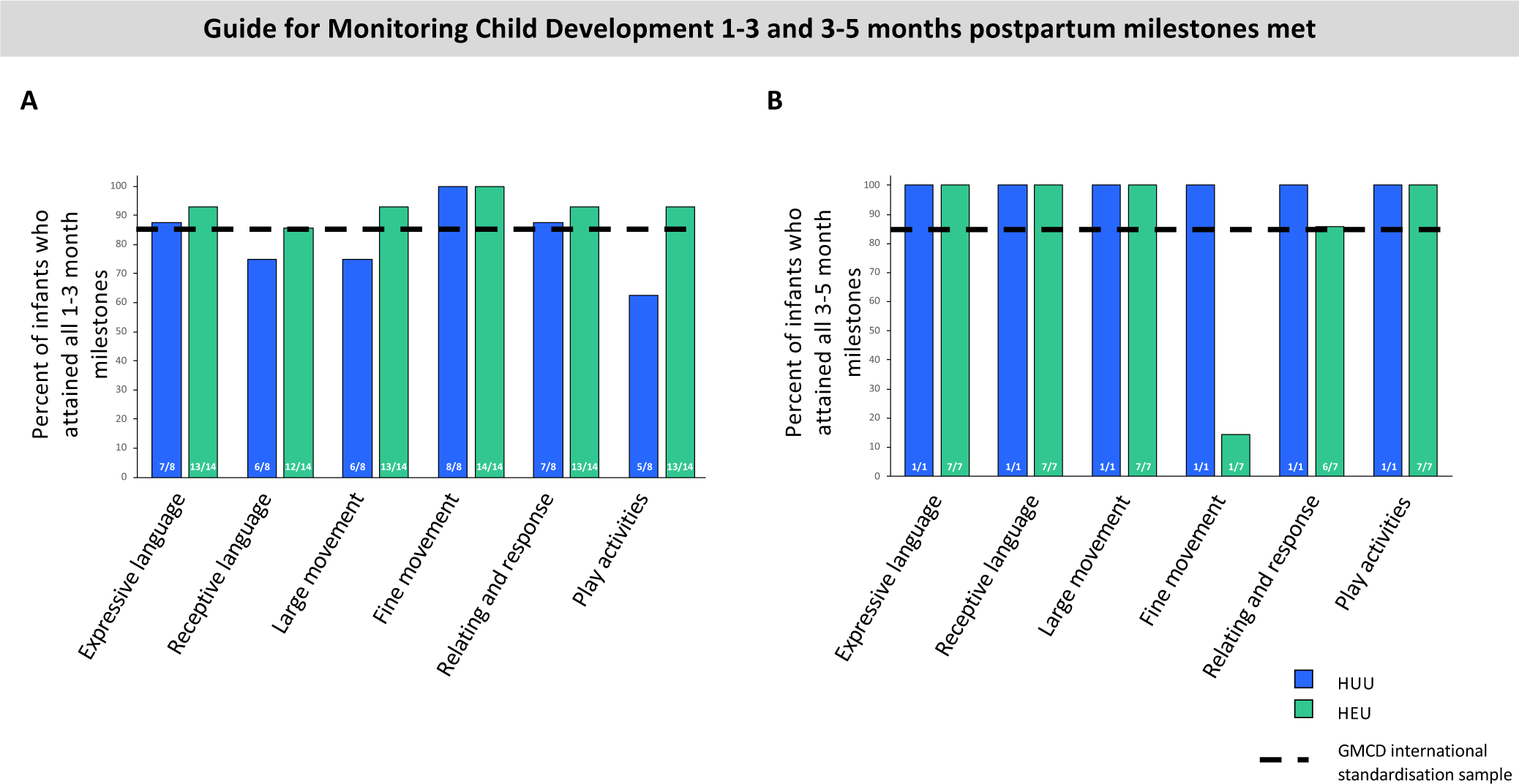
Relationships between in utero HIV-exposure and attainment of GMCD neurodevelopmental milestones 1-3 months (A) and 3-5 months (B) of age. There was no detected difference in the proportion of infants who were HUU (n=8) compared to HEU (n=14) who attained GMCD milestones at 1-3 months of age ([p>0.05], Fisher’s exact), and no comparisons between infants who were HUU vs. HEU were made for 3-5 month milestones met, given that only 1/10 infants who were HUU at follow up was 3-5 months of age. All infants met milestones when comparing to the GMCD standardisation sample, except for infants who were HUU for 1-3 month receptive language, large movement, and play activities milestones, and infants who were HEU for fine movement milestones at 3-5 months of age. Bar graphs are proportion (%) attaining all age-appropriate GMCD milestones. GMCD = Guide for monitoring child development; HUU = HIV-unexposed, uninfected; HEU = HIV-exposed, uninfected.

A 3-5-month assessment was performed on the 8/31 (25.8%) infants at follow up who were within this age range. Of these infants, 7/8 (87.5%) were HEU. Fewer infants at 3-5 months of age who were HEU had attained age-appropriate fine movement milestones in comparison to the GMCD international standardised sample (14.3% [n=1/7] of HEU attained vs. 85% standardised sample, Fig 4B). All of these infants who were HEU had attained all age-appropriate milestones for expressive language, receptive language, large movement and play activities, and met the international standardised sample proportion for relating and response behaviour (+0.70%). The one infant who was HUU and 3-5 months of age attained all developmental milestones.

### Differences between the cohort at birth and 12 weeks postpartum

Within the group of infants who were HUU, those who were not lost to follow up had lower BMI at birth than those who were lost to follow up (-1.32 ± 2.47 vs. 0.90 ± 1.85, p=0.03). There were no other differences for anthropometric, Apgar scores or immune measures at birth within the HUU or HEU infant groups, when comparing infant who were versus were not at follow up.

### Exploring food security and maternal nutrition

#### Food security and nutrient intakes among women living with and without HIV

There were no differences between mothers living with and without HIV for probability of reporting household food insecurity or DDS (Table 3). Mothers living with HIV had higher intakes of Vitamin D (64.5 [42.0, 84.6] vs. 8.60 [0.38, 20.8], p=0.002) and Se (51.7 [42.1, 73.7] vs. 12.6 [7.41, 34.4], p<.001) compared to mothers living without HIV (Table 3). Full data on absolute nutrient intake levels for mothers living with and without HIV from the 24-hour dietary recall are presented in Supplementary table S3.

**Table 3.** Maternal reports of household food security and nutrient intakes from one 24-hour dietary recall for mothers with and without HIV who attended follow up.

|  | Total<br>N=31 | HIV-uninfected<br>(n=10) | HIV-infected<br>(n=21) | p value |
| --- | --- | --- | --- | --- |
| **Household food security circumstances (n)** |  |  |  |  |
| Maternal reports of worrying about food runout |  |  |  | NS |
| Never occurs | 14 | 3 | 11 |  |
| Occurs often/sometimes | 17 | 7 | 10 |  |
| Maternal reports of experiencing about food runout |  |  |  | NS |
| Never occurs | 16 | 4 | 12 |  |
| Occurs often/sometimes | 15 | 6 | 9 |  |
| Maternal reports of being unable to afford balanced meals |  |  |  | NS |
| Never occurs | 9 | 2 | 7 |  |
| Occurs often/sometimes | 26 | 8 | 14 |  |
| **Maternal nutrient intakes** |  |  |  |  |
| Dietary diversity score (/9) | 4.00 (4.00, 6.00) | 4.00 (3.00, 5.25) | 4.00 (3.00, 6.00) | NS |
| Dietary diversity score <4 (n) | 7 | 3 | 4 | NS |
| Median intake (%) of TULs by mothers |  |  |  |  |
| *Minerals* |  |  |  |  |
| Ca | 5.48 (3.40, 9.56) | 7.78 (4.09, 14.4) | 5.28 (3.40, 7.48) | NS |
| Fe | 24.7 (16.7, 27.8) | 25.0 (15.5, 30.1) | 24.2 (16.4, 27.8) | NS |
| Mg | 52.9 (34.6, 81.1) | 80.0 (33.0, 99.1) | 48.0 (38.4, 64.0) | NS |
| P | 14.9 (10.8, 19.3) | 18.1 (10.4, 21.4) | 14.3 (10.5, 18.3) | NS |
| Na | 69.9 (48.2, 154) | 92.4 (43.9, 171) | 69.9 (47.8, 146) | NS |
| Zn | 20.9 (16.8, 29.3) | 17.4 (15.2, 23.8) | 23.7 (19.8, 29.9) | NS |
| Cu | 7.70 (5.50, 11.0) | 11.4 (4.97, 13.4) | 7.40 (5.20, 9.55) | NS |
| Se | 7.00 (4.15, 9.78) | 1.86 (1.09, 5.08) | 7.63 (6.21, 10.9) | <.001 |
| Mn | 12.7 (7.52, 17.6) | 15.7 (6.64, 21.9) | 10.2 (7.28, 16.2) | NS |
| I | 8.36 (4.09, 16.0) | 8.27 (2.30, 16.1) | 8.36 (5.86, 17.2) | NS |
| *Vitamins* |  |  |  |  |
| Vitamin A | 14.1 (10.2, 23.8) | 14.1 (11.1, 24.2) | 14.3 (10.0, 23.5) | NS |
| Niacin | 44.0 (30.6, 55.7) | 41.6 (29.1, 57.9) | 46.6 (28.5, 55.9) | NS |
| Vitamin B6 | 2.88 (1.60, 3.80) | 2.61 (1.37, 5.19) | 3.13 (1.72, 3.72) | NS |
| Folate | 25.9 (20.5, 31.9) | 23.9 (18.3, 38.3) | 27.8 (20.7, 31.2) | NS |
| Vitamin C | 1.15 (0.40, 3.65) | 1.58 (0.11, 4.74) | 1.10 (0.53, 3.13) | NS |
| Vitamin D | 4.29 (0.83, 7.86) | 0.86 (0.04, 2.08) | 6.45 (4.20, 8.46) | 0.002 |
| Vitamin E | 0.68 (0.44, 1.19) | 0.52 (0.33, 1.17) | 0.80 (0.47, 1.33) | NS |
| **Median intake (%) of EARs by mothers** |  |  |  |  |
| *Macronutrients* |  |  |  |  |
| Protein | 66.6 (49.9, 90.4) | 68.7 (60.6, 87.7) | 65.5 (48.9, 99.5) | NS |
| Carbohydrates | 98.7 (63.6, 113) | 81.8 (56.7, 81.8) | 99.8 (77.7, 118) | NS |
| *Vitamins* |  |  |  |  |
| Vitamin A | 47.0 (34.1, 75.4) | 46.9 (36.8, 77.6) | 47.8 (33.4, 78.3) | NS |
| Thiamin | 90.0 (56.7, 111) | 88.3 (53.5, 116) | 90.0 (57.5, 118) | NS |
| Riboflavin | 55.4 (43.1, 74.6) | 38.9 (24.4, 96.9) | 58.5 (49.6, 74.2) | NS |
| Niacin | 119 (72.3, 150) | 112 (69.4, 156) | 125 (75.4, 150) | NS |
| Vitamin B6 | 170 (88.7, 223) | 139 (80.6, 306) | 184 (97.4, 219) | NS |
| Folate | 54.2 (43.1, 70.9) | 47.1 (39.9, 85.1) | 61.8 (45.9, 69.2) | NS |
| Vitamin B12 | 70.8 (62.5, 108) | 68.8 (9.38, 102) | 75.0 (66.7, 133) | NS |
| Vitamin C | 23.0 (8.00, 73.0) | 31.5 (2.25, 92.7) | 22.0 (10.5, 62.5) | NS |
| Vitamin D | 42.9 (8.30, 78.6) | 8.60 (0.38, 20.8) | 64.5 (42.0, 84.6) | 0.002 |
| Vitamin E | 42.5 (27.4, 74.3) | 32.7 (20.8, 73.1) | 49.4 (29.5, 82.8) | NS |
| *Minerals* |  |  |  |  |
| Ca | 17.1 (10.6, 29.9) | 24.3 (12.7, 44.5) | 16.5 (10.6, 23.4) | NS |
| Fe | 171 (115, 192) | 168 (112, 192) | 173 (106, 208) | NS |
| Mg | 71.3 (45.7, 111) | 110 (44.0, 136) | 63.4 (50.8, 87.1) | NS |
| P | 103 (68.3, 133) | 125 (41.2, 147) | 98.8 (69.5, 126) | NS |
| Zn | 78.9 (64.5, 113) | 66.8 (47.4, 91.6) | 91.0 (73.3, 115) | NS |
| Cu | 77.0 (52.8, 110) | 112 (48.1, 127) | 74.0 (50.4, 95.5) | NS |
| Se | 47.5 (28.1, 66.3) | 12.6 (7.41, 34.4) | 51.7 (42.1, 73.7) | <.001 |
| I | 44.0 (21.5, 80.9) | 43.5 (12.1, 80.4) | 44.0 (30.4, 90.4) | NS |

Data from mother-infant dyads that attended follow up are presented as median (IQR) and p values are from one-way analysis of variance (Kruskal-Wallis/Wilcoxon test for non-parametric data; ANOVA for normal data with equal variance). Dietary diversity score was calculated using nine food groups, only counting each food groups once. Percent intake of TULs and EARs were calculated using the Institute of Medicine's TULs for minerals and vitamins for lactating women 14-18, 19-30 or 31-50 years of age<sup>26</sup>. Iodised salt was assumed to be consumed by all mothers, as the majority of salt consumed in South Africa is iodised<sup>64</sup>. TULs = Tolerable upper levels; EARs = Estimated average requirements.

#### Household food insecurity and diet quality

Full data on maternal reports of food insecurity and nutrient intakes is presented in Supplementary table S4. Overall, a large proportion of mothers were at risk of inadequate intake of macronutrients, vitamins, and minerals, irrespective of reports of worrying about (Supplementary fig 3) or experiencing (Supplementary fig 4) food runout, or inability to afford balanced meals (Supplementary fig 5). Food insecurity was associated with an increased risk of inadequate intake (median %EAR met) of vitamin B12 amongst mothers who reported experiencing (66.7 [12.5, 91.7] vs. 89.6 [67.7, 144], p=0.01 [Supplementary fig S4]) food runout, or an inability to afford balanced meals (66.7 [19.8, 96.9] vs. 108 [70.8, 150], p=0.04 [Supplementary fig S5]) compared to mothers who did not. Of the mothers who reported worrying about or experiencing food runout, or inability to afford balanced meals, 76.5%, 86.7%, and 81.8% were at risk for inadequate intake, respectively. Overall, very few mothers, irrespective of food insecurity reports, had intake of vitamins or minerals that was too high, however, TULs were exceeded for magnesium and sodium (Supplementary table S4). There were no relationships between reports of household food insecurity and DDS (Supplementary table S4).

**Figure 5.**
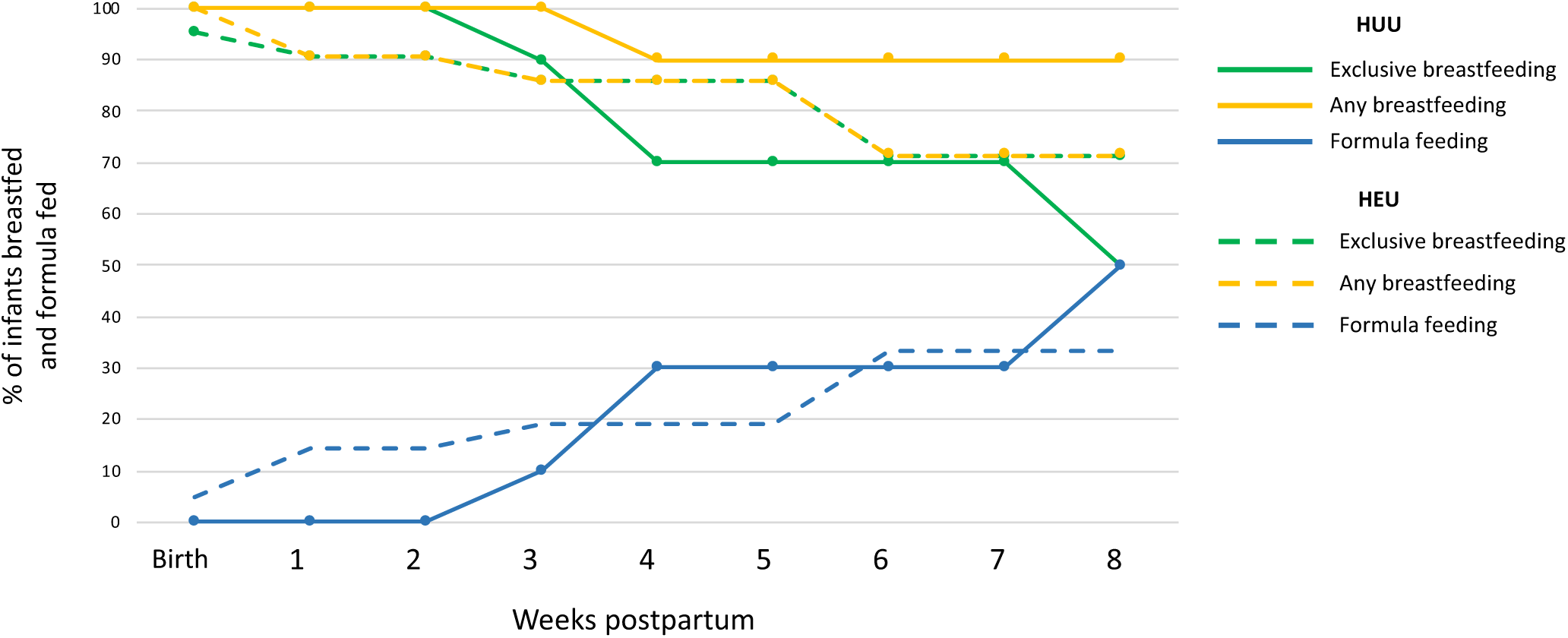
Feeding patterns from birth to 8 weeks postpartum for infants who were HUU and HEU. There were no differences in the likelihood of being exclusively breastfed (EBF) at 12 weeks of age for infants who were HUU (n=10) compared to HEU (n=21; [p>0.05], Fisher’s exact 2-Tail). Data on feeding practices were available for the whole cohort from birth to 8 weeks postpartum. Each point on the line represents the proportion (%) of infants who were HUU or HEU and were exclusively breastfed, receiving any breastmilk, or were formula fed at that time (weeks). HUU = HIV-unexposed, uninfected; HEU = HIV-exposed, uninfected.

#### Infant feeding patterns

To explore the feeding patterns of infants who were HEU compared to HUU, we plotted the percentage of infants who were HEU or HUU and were breastfed (mixed and exclusive) or formula fed from birth to 8 weeks of age (Fig 5). Feeding practices were only available until 8 weeks of age for the youngest infant at follow up, so this was chosen at the cut off to plot feeding patterns for the whole pilot cohort. At birth, exclusive breastfeeding (EBF) was initiated for all but one infant, who was HEU and received mixed feeds. At follow up, slightly more infants who were HEU were still being exclusively breastfed compared to HUU, however, most infants who were HUU were still receiving some breastmilk (Fig 5). There were no differences between infants who were HUU compared to HEU for likelihood of EBF at follow up (Table 2). Mothers who reported never experiencing food runout or were always able to afford balanced meals were more likely than mothers who experienced food runout, or were unable to afford balanced meals, to be EBF at follow up, irrespective of maternal HIV status (Table 4).

**Table 4.**
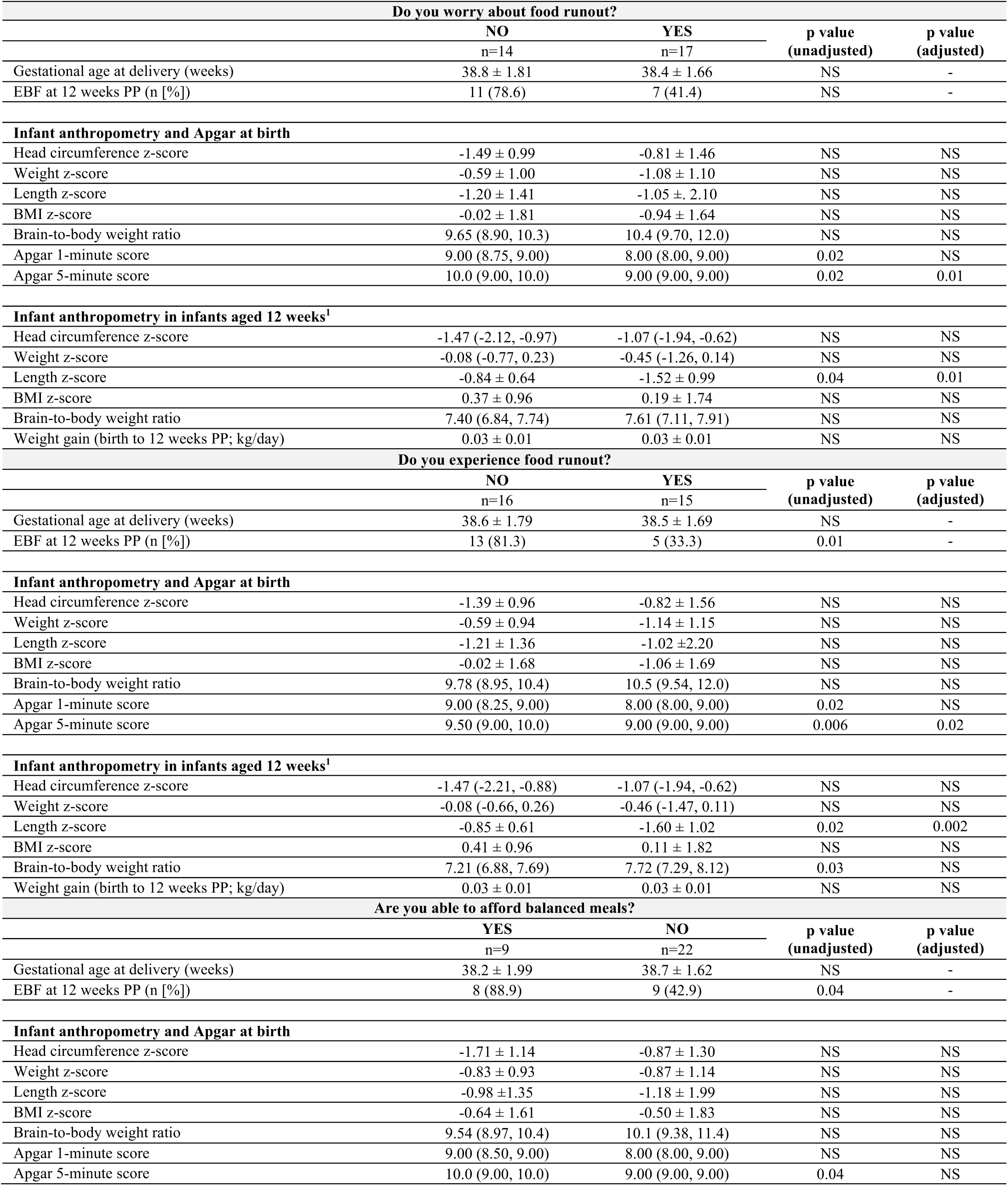
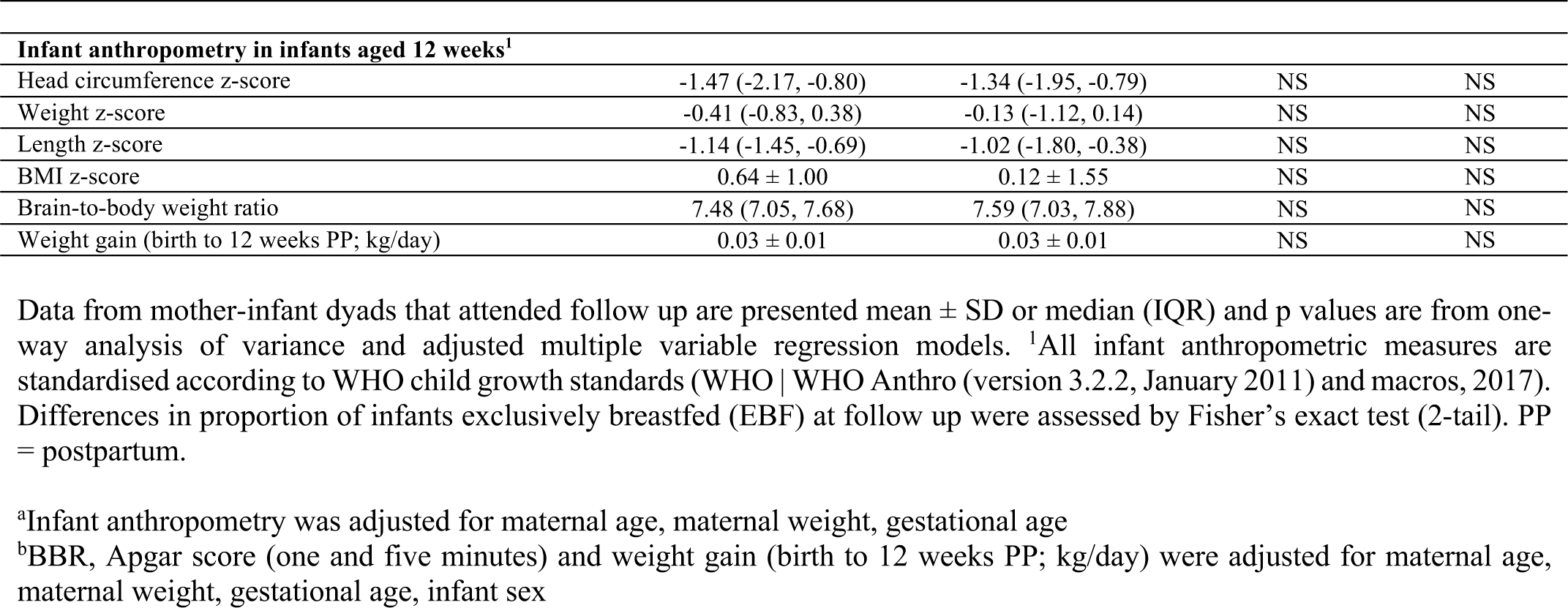
Anthropometry and Apgar scores at birth, and anthropometry and breastfeeding practices at 12 weeks postpartum for infants exposed to food-insecure conditions compared to those who were not.

#### Influence of food insecurity on infant Apgar scores and growth outcomes at birth

Maternal reports of food insecurity did not appear to influence infant gestational age or infant anthropometry at birth (Table 4). Apgar score at five minutes was slightly lower for infants whose mothers reported worrying about (9.00 [9.00, 9.00] vs. 10.0 [9.00, 10.0], p=0.01) or experiencing (9.00 [9.00, 9.00] vs. 9.50 [9.00, 10.0], p=0.02) food runout compared to those who reported not worrying about or experiencing food runout (Table 4).

#### Influence of food insecurity on infant growth outcomes at 12 weeks

Infant length at 12 weeks was lower among infants whose mothers reported worrying about (-1.52 ± 0.99 vs. -0.84 ± 0.64, p=0.01) and experiencing (-1.60 ± 1.02 vs. -0.85 ± 0.61, p=0.002) food runout in comparison to those who did not (Table 4). There were no other effects of food security on infant anthropometry at 12 weeks (Table 4).

#### HEU may increase vulnerability to effects of food insecurity on risk of stunting at birth

Among infants whose mothers reported worrying about food runout, infants who were HEU had increased risk of stunting at birth compared to infants who were HUU (p=0.04, Fisher’s exact test [2-tail]; [Supplementary fig 6A]). Maternal HIV status did not influence the relationship between maternal reports of food insecurity and any other infant outcomes at birth or 12 weeks postpartum.

#### Influence of food insecurity on neurodevelopmental outcomes in 12 week old infants

The probability of attaining all 1-3 month GMCD expressive and receptive language, large movement, play activities and relating and response behaviour milestones did not associate with maternal reports of household food insecurity (Supplementary fig S7). No comparisons were made for fine movement outcomes between infants exposed to food insecure conditions compare to those who were not, as all infants who were 1-3 months of age at follow up met all age-appropriate fine movement milestones. At 3-5 months, there were no differences in the proportion of infants who attained fine movement and relating and response behaviour milestones based on maternal reports of food security (Supplementary fig S7). No comparisons were made for expressive and receptive language, large movement, or play activities, as all infants 3-5 months of age at follow up had met these milestones. It was not possible to further stratify these comparisons based on maternal HIV status due to the small sample size of the pilot.

## Discussion

As access to ART increases worldwide and the number of infants born each year who are HIV-exposed but uninfected also rises, there is growing need to understand how, and to what extent, exposure to HIV and ART *in utero* and during breastfeeding influences infant health trajectories. In this small pilot study, we recruited a cohort of women living with and without HIV to test the feasibility of carrying out a full-scale, observational study to investigate the effects of HEU on growth, immune- and neuro-development in infants in early life. Our hypothesis-generating analyses revealed that infants who are HEU may have reduced head circumference and elevated CCR2 expression by CD14^+^ monocytes within four days of birth compared to infants who are HUU. Our exploratory analyses also suggest that food insecurities, and the likely ensuing poor maternal nutritional status, may adversely affect the growth and neurodevelopment of infants in the first four months of life, and at least for some measures, the effects of a suboptimal early life nutritional environment may be most detrimental for infants who are HEU. While these exploratory analyses are underpowered to make conclusive statements, findings will inform our research questions and analyses in our full-scale, observational study aimed at better understanding these exposure-outcome relationships.

Our finding that infants who are HEU may have reduced head circumference at birth compared to infants who are HUU is in agreement with other studies, which found that HEU associates with lower weight, length, BMI^40,41,^ and head circumference^42^ at birth compared to infants who are HUU. Small head circumference at birth has been shown to associate with poorer performance on neurodevelopmental assessments in school-aged children, including on cognitive tasks measuring memory and visuo-spatial ability^43^, early adiposity rebound and increased risk for adult obesity^44,45^, cardiovascular disease mortality^46^, and mental health disorders such as schizophrenia^47^. It is not known whether, in the context of maternal HIV infection, small head circumference at birth is linked to persistent deficits in neurodevelopment and/or, in the longer term, later life brain and metabolic compromise. However, the effect of HEU on neurodevelopment and growth outcomes will be further investigated in our full-scale study over the first two-years.

We also observed elevated CCR2 expression on classical monocytes at birth, and increased levels of CCR2 expression (MFI) on intermediate monocytes at birth and 12 weeks in infants who were HEU. Increased CCR2 expression may result in increased recruitment of monocyte populations across the blood brain barrier, which may have consequences for neurodevelopment, as the expression of CCR2 by CD14^+^ monocytes has been shown to associate with HIV-1 induced neuropsychological impairment and neuroinflammation in adults^20,48^. The expression and release of a CCR2 ligand, monocyte chemoattractant protein-1 (MCP-1), from astrocytes in the brain is increased by HIV-1 infection^49^. MCP-1 levels have been shown to positively correlate with severity of HIV-induced neuropsychological impairment in adults^50^. Importantly, whether or not elevated MCP-1 levels or CCR2 expression by monocyte subsets have consequences for children perinatally exposed to HIV remains to be determined and will be examined further in our scaled-up cohort.

When exploring relationships between food security and infant outcomes, we found that household food insecurity may associate with reduced infant length at 12 weeks postpartum, and infants who also experience HEU may be at an increased risk of stunting at birth compared to infants who are HUU and whose mothers also experience food insecure conditions. Stunting is the most common manifestation of infant undernutrition globally^51^. Our findings may suggest that infants who are HEU are distinctly vulnerable to the programming effects of suboptimal nutrition *in utero* and postnatally, and future studies should further investigate the mechanisms that may underly this relationship.

Differences in maternal nutrient intakes between mothers living with and without HIV were minimal, and mothers were at risk for inadequate macronutrient intakes irrespective of food insecurity reports. We also found maternal reports of experiencing food insecurity associated with lower vitamin B12, and a large proportion of mothers, irrespective of food insecurity circumstances, were at risk for inadequate intakes. Inadequate maternal vitamin B12 intakes have shown to cause secondary deficiencies in breastfed infants in the first six months of life, leading to delayed growth and neurodevelopment^52^. Food insecurity is also a known barrier to exclusive breastfeeding^53^, and mothers experiencing food insecurity may be more likely to return to work soon after birth^54^, or may have challenges maintaining milk supply due to inadequate nutrition^55^. In agreement with this, we found that mothers who never experienced food runout or were always able to afford balanced meals were more likely to be exclusively breastfeeding at follow up. Given the high rates of reported food insecurity among the pilot study cohort and the potential influence of food and nutrition insecurity on maternal nutrient intakes, we will be including these important variables in our full-scale study, which will allow us to assess relationships between food and nutrition insecurity, maternal HIV infection, and infant feeding practices and development over a 24-month period, encompassing the recommended EBF (six months) and mixed feeding (24 months) periods^56^.

Importantly, the translatability of the exploratory analyses presented are limited by the pilot’s small sample size, as these analyses were intended to inform the development and refinement of our research questions and study protocol for a full-scale study investigating relationships between the early life nutritional environment, HEU and infant development. There was high attrition amongst both populations, particularly within the HUU group, which may have led to sample bias at follow up. Individuals from communities in this area are highly mobile, and although multiple attempts were made to contact each mother to encourage her to return, this presented challenges for study retention. Despite these limitations, our study was able to capture information on infant growth, immune function and neurodevelopment at two time points within the same infant population. We are currently conducting a larger prospective pregnancy and birth cohort study at Kalafong Hospital to further investigate relationships that have emerged in these exploratory pilot study analyses. In an aim to improve participant retention in the full-scale study, Ward-based Primary Health Care Outreach teams have been employed to trace and contact women who miss a follow up appointment. To our knowledge, this prospective cohort study is among the few to concomitantly interrogate growth and neurodevelopmental outcomes, immune function, and maternal nutrition and food security among a population of infants who are HEU in comparison to infants who are HUU.

## Conclusions

Study participant retention was challenging in this pilot, however, the study helped to identify barriers to recruitment and retention that were used to inform a revised full-scale study protocol. Exploratory data analyses revealed possible relationships between exposure to maternal HIV infection in the womb, household food insecurity, and infant outcomes at birth and 12 weeks postpartum. We are now investigating these relationships in a full-scale, longitudinal observational study.

## Data Availability

The datasets generated during and/or analysed during the current study are available from the corresponding author on reasonable request.

## Acknowledgements

The authors would like to thank the participating families and health care workers in Pretoria, for without whom this research would not be possible. The researchers thank the Obstetric and Paediatric services and management of Kalafong Provincial Tertiary Hospital for their support. Sister Sinah Lebogo and Dr Kerstin Bartelheim are acknowledged for the patient recruitment and data capturing. The researchers also thank Ms Gisela van Dyk for the processing of samples.

---

**Supplementary table S1.** Flow cytometry reagent list (including lasers and detectors used).

| Monoclonal antibody | Clone | Supplier | Excitation (laser) | Fluorescent Channel | Emission (Detector) |
| --- | --- | --- | --- | --- | --- |
| 7-AAD | N/A | Beckman Coulter, Miami, FL, USA | 488 nm (blue) | FL4 | 695/30 |
| CD16-FITC | 3G8 | BioLegend®, San Diego, CA, USA | 488 nm (blue) | FL1 | 525/40 |
| CD14-APC | 63D3 | BioLegend®, San Diego, CA, USA | 635 nm (red) | FL6 | 660/20 |
| CD192 (CCR2)-PE | K036C2 | BioLegend®, San Diego, CA, USA | 488 nm (blue) | FL2 | 575/30 |

N/A: Not applicable

**Supplementary table S2.** Flow cytometry compensation matrix.

| % Spillover |  |  |  |  |
| --- | --- | --- | --- | --- |
|  | FL1 | FL2 | FL4 | FL6 |
| FL1 | - | 2.40 | 0.00 | 0.00 |
| FL2 | 5.00 | - | 0.00 | 0.00 |
| FL4 | 0.00 | 59.98 | - | 0.00 |
| FL6 | 0.00 | 0.00 | 0.00 | - |

**Supplementary table S3.**
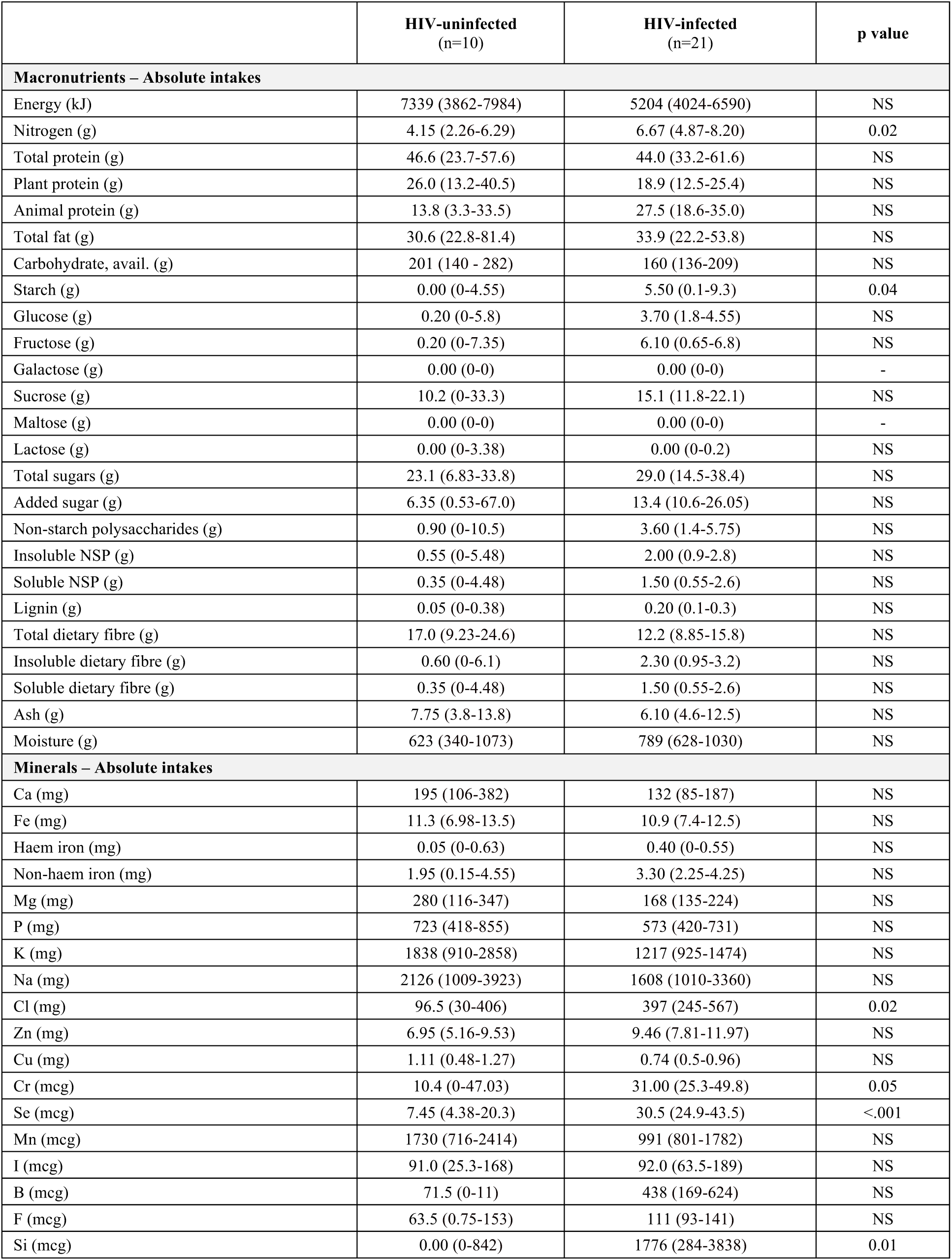
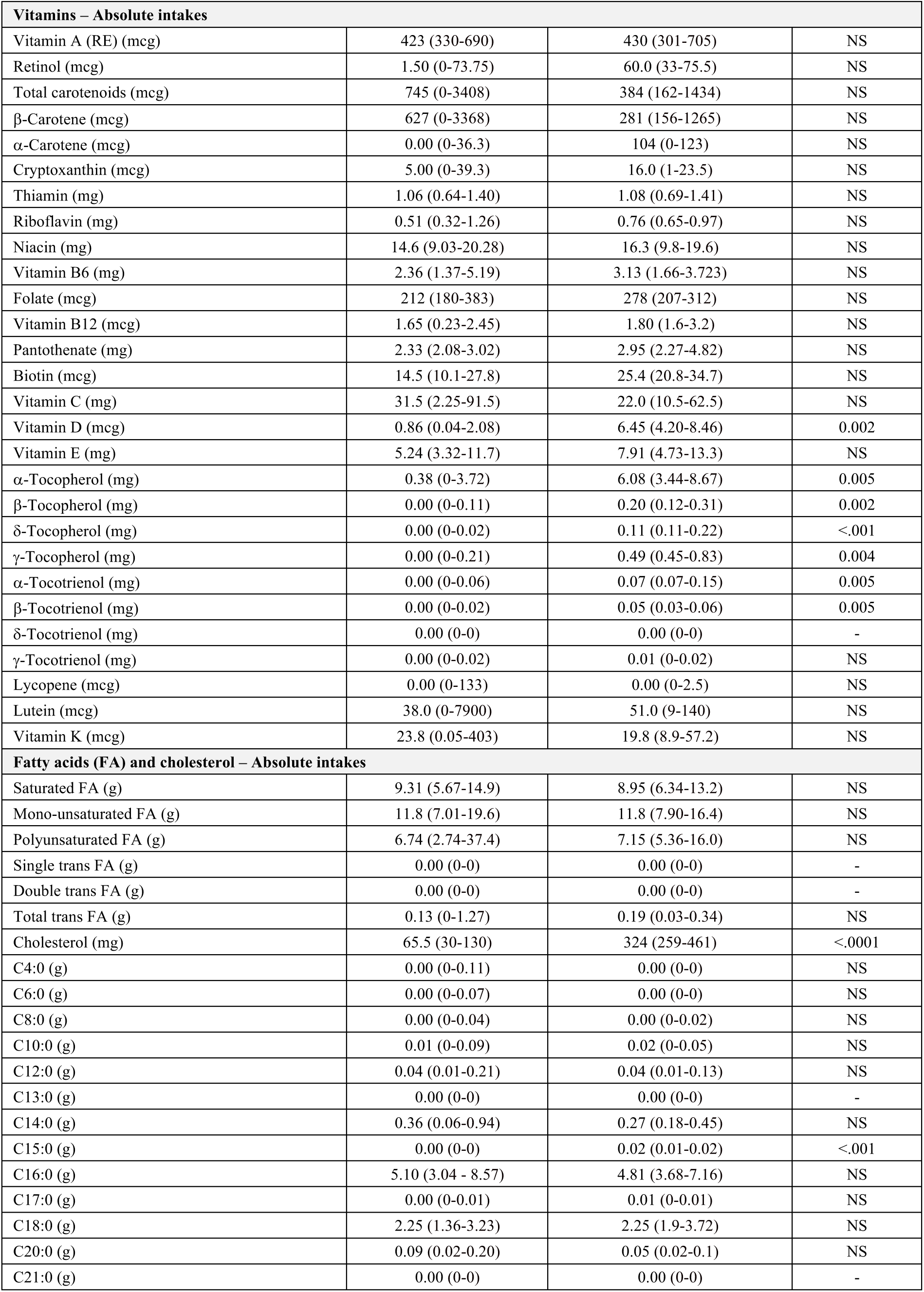
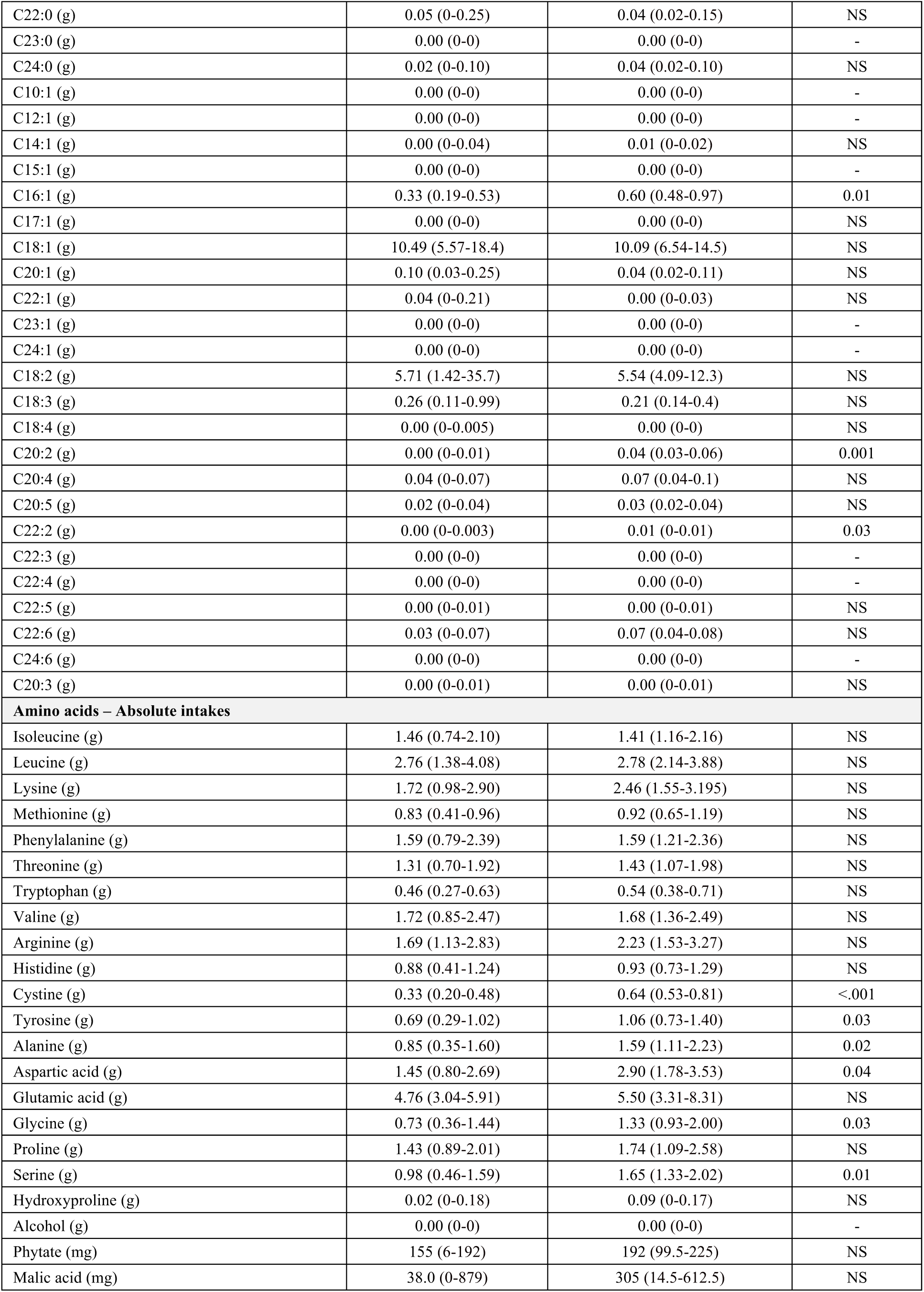
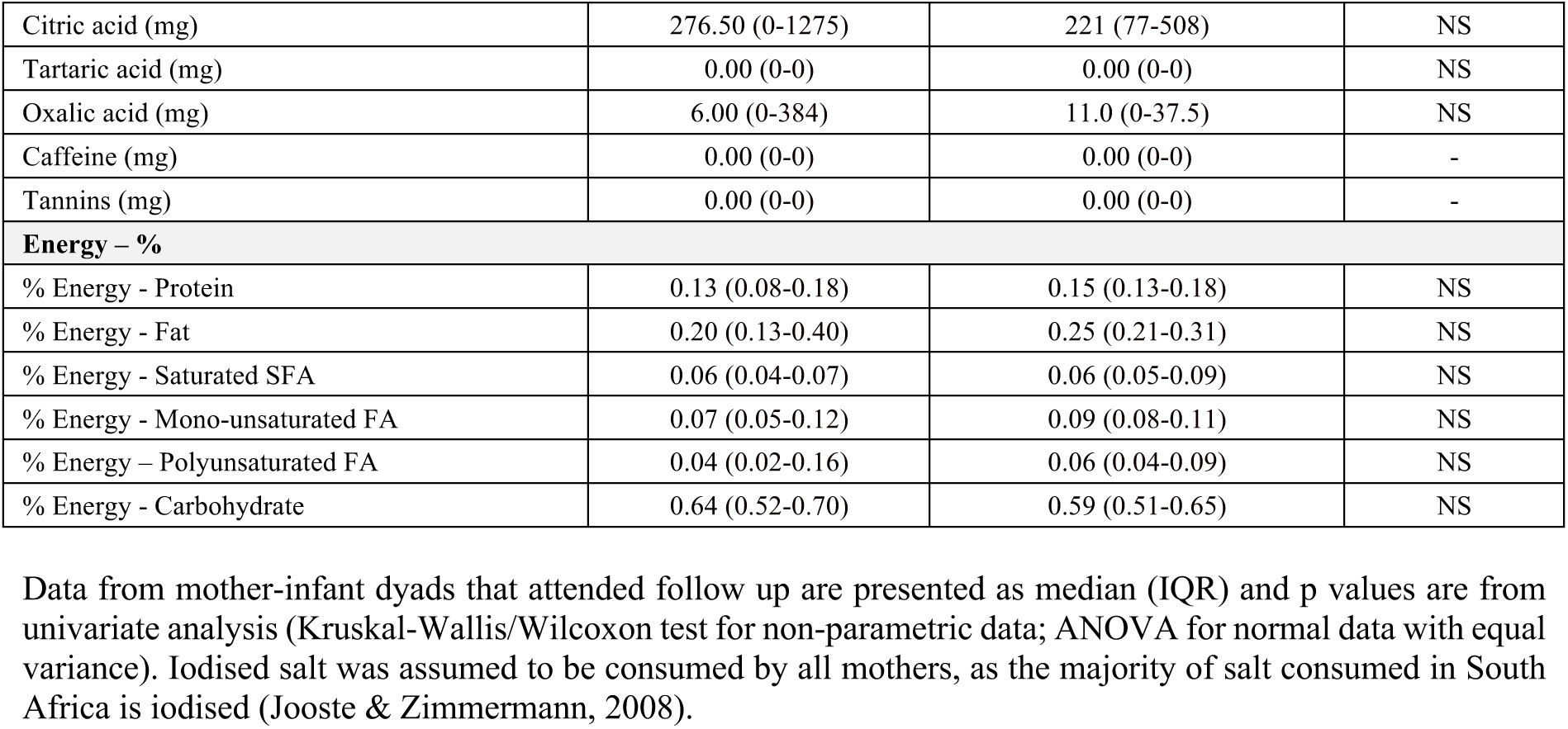
Maternal nutrient intakes from one 24-hour dietary recall for mothers with and without HIV who attended follow up.

|  | HIV-uninfected<br>(n=10) | HIV-infected<br>(n=21) | p value |
| --- | --- | --- | --- |
| **Macronutrients – Absolute intakes** |  |  |  |
| Energy (kJ) | 7339 (3862-7984) | 5204 (4024-6590) | NS |
| Nitrogen (g) | 4.15 (2.26-6.29) | 6.67 (4.87-8.20) | 0.02 |
| Total protein (g) | 46.6 (23.7-57.6) | 44.0 (33.2-61.6) | NS |
| Plant protein (g) | 26.0 (13.2-40.5) | 18.9 (12.5-25.4) | NS |
| Animal protein (g) | 13.8 (3.3-33.5) | 27.5 (18.6-35.0) | NS |
| Total fat (g) | 30.6 (22.8-81.4) | 33.9 (22.2-53.8) | NS |
| Carbohydrate, avail. (g) | 201 (140 - 282) | 160 (136-209) | NS |
| Starch (g) | 0.00 (0-4.55) | 5.50 (0.1-9.3) | 0.04 |
| Glucose (g) | 0.20 (0-5.8) | 3.70 (1.8-4.55) | NS |
| Fructose (g) | 0.20 (0-7.35) | 6.10 (0.65-6.8) | NS |
| Galactose (g) | 0.00 (0-0) | 0.00 (0-0) | - |
| Sucrose (g) | 10.2 (0-33.3) | 15.1 (11.8-22.1) | NS |
| Maltose (g) | 0.00 (0-0) | 0.00 (0-0) | - |
| Lactose (g) | 0.00 (0-3.38) | 0.00 (0-0.2) | NS |
| Total sugars (g) | 23.1 (6.83-33.8) | 29.0 (14.5-38.4) | NS |
| Added sugar (g) | 6.35 (0.53-67.0) | 13.4 (10.6-26.05) | NS |
| Non-starch polysaccharides (g) | 0.90 (0-10.5) | 3.60 (1.4-5.75) | NS |
| Insoluble NSP (g) | 0.55 (0-5.48) | 2.00 (0.9-2.8) | NS |
| Soluble NSP (g) | 0.35 (0-4.48) | 1.50 (0.55-2.6) | NS |
| Lignin (g) | 0.05 (0-0.38) | 0.20 (0.1-0.3) | NS |
| Total dietary fibre (g) | 17.0 (9.23-24.6) | 12.2 (8.85-15.8) | NS |
| Insoluble dietary fibre (g) | 0.60 (0-6.1) | 2.30 (0.95-3.2) | NS |
| Soluble dietary fibre (g) | 0.35 (0-4.48) | 1.50 (0.55-2.6) | NS |
| Ash (g) | 7.75 (3.8-13.8) | 6.10 (4.6-12.5) | NS |
| Moisture (g) | 623 (340-1073) | 789 (628-1030) | NS |
| **Minerals – Absolute intakes** |  |  |  |
| Ca (mg) | 195 (106-382) | 132 (85-187) | NS |
| Fe (mg) | 11.3 (6.98-13.5) | 10.9 (7.4-12.5) | NS |
| Haem iron (mg) | 0.05 (0-0.63) | 0.40 (0-0.55) | NS |
| Non-haem iron (mg) | 1.95 (0.15-4.55) | 3.30 (2.25-4.25) | NS |
| Mg (mg) | 280 (116-347) | 168 (135-224) | NS |
| P (mg) | 723 (418-855) | 573 (420-731) | NS |
| K (mg) | 1838 (910-2858) | 1217 (925-1474) | NS |
| Na (mg) | 2126 (1009-3923) | 1608 (1010-3360) | NS |
| Cl (mg) | 96.5 (30-406) | 397 (245-567) | 0.02 |
| Zn (mg) | 6.95 (5.16-9.53) | 9.46 (7.81-11.97) | NS |
| Cu (mg) | 1.11 (0.48-1.27) | 0.74 (0.5-0.96) | NS |
| Cr (mcg) | 10.4 (0-47.03) | 31.00 (25.3-49.8) | 0.05 |
| Se (mcg) | 7.45 (4.38-20.3) | 30.5 (24.9-43.5) | <.001 |
| Mn (mcg) | 1730 (716-2414) | 991 (801-1782) | NS |
| I (mcg) | 91.0 (25.3-168) | 92.0 (63.5-189) | NS |
| B (mcg) | 71.5 (0-11) | 438 (169-624) | NS |
| F (mcg) | 63.5 (0.75-153) | 111 (93-141) | NS |
| Si (mcg) | 0.00 (0-842) | 1776 (284-3838) | 0.01 |

| **Vitamins – Absolute intakes** |  |  |  |
| --- | --- | --- | --- |
| Vitamin A (RE) (mcg) | 423 (330-690) | 430 (301-705) | NS |
| Retinol (mcg) | 1.50 (0-73.75) | 60.0 (33-75.5) | NS |
| Total carotenoids (mcg) | 745 (0-3408) | 384 (162-1434) | NS |
| β-Carotene (mcg) | 627 (0-3368) | 281 (156-1265) | NS |
| α-Carotene (mcg) | 0.00 (0-36.3) | 104 (0-123) | NS |
| Cryptoxanthin (mcg) | 5.00 (0-39.3) | 16.0 (1-23.5) | NS |
| Thiamin (mg) | 1.06 (0.64-1.40) | 1.08 (0.69-1.41) | NS |
| Riboflavin (mg) | 0.51 (0.32-1.26) | 0.76 (0.65-0.97) | NS |
| Niacin (mg) | 14.6 (9.03-20.28) | 16.3 (9.8-19.6) | NS |
| Vitamin B6 (mg) | 2.36 (1.37-5.19) | 3.13 (1.66-3.723) | NS |
| Folate (mcg) | 212 (180-383) | 278 (207-312) | NS |
| Vitamin B12 (mcg) | 1.65 (0.23-2.45) | 1.80 (1.6-3.2) | NS |
| Pantothenate (mg) | 2.33 (2.08-3.02) | 2.95 (2.27-4.82) | NS |
| Biotin (mcg) | 14.5 (10.1-27.8) | 25.4 (20.8-34.7) | NS |
| Vitamin C (mg) | 31.5 (2.25-91.5) | 22.0 (10.5-62.5) | NS |
| Vitamin D (mcg) | 0.86 (0.04-2.08) | 6.45 (4.20-8.46) | 0.002 |
| Vitamin E (mg) | 5.24 (3.32-11.7) | 7.91 (4.73-13.3) | NS |
| α-Tocopherol (mg) | 0.38 (0-3.72) | 6.08 (3.44-8.67) | 0.005 |
| β-Tocopherol (mg) | 0.00 (0-0.11) | 0.20 (0.12-0.31) | 0.002 |
| δ-Tocopherol (mg) | 0.00 (0-0.02) | 0.11 (0.11-0.22) | <.001 |
| γ-Tocopherol (mg) | 0.00 (0-0.21) | 0.49 (0.45-0.83) | 0.004 |
| α-Tocotrienol (mg) | 0.00 (0-0.06) | 0.07 (0.07-0.15) | 0.005 |
| β-Tocotrienol (mg) | 0.00 (0-0.02) | 0.05 (0.03-0.06) | 0.005 |
| δ-Tocotrienol (mg) | 0.00 (0-0) | 0.00 (0-0) | - |
| γ-Tocotrienol (mg) | 0.00 (0-0.02) | 0.01 (0-0.02) | NS |
| Lycopene (mcg) | 0.00 (0-133) | 0.00 (0-2.5) | NS |
| Lutein (mcg) | 38.0 (0-7900) | 51.0 (9-140) | NS |
| Vitamin K (mcg) | 23.8 (0.05-403) | 19.8 (8.9-57.2) | NS |
| **Fatty acids (FA) and cholesterol – Absolute intakes** |  |  |  |
| Saturated FA (g) | 9.31 (5.67-14.9) | 8.95 (6.34-13.2) | NS |
| Mono-unsaturated FA (g) | 11.8 (7.01-19.6) | 11.8 (7.90-16.4) | NS |
| Polyunsaturated FA (g) | 6.74 (2.74-37.4) | 7.15 (5.36-16.0) | NS |
| Single trans FA (g) | 0.00 (0-0) | 0.00 (0-0) | - |
| Double trans FA (g) | 0.00 (0-0) | 0.00 (0-0) | - |
| Total trans FA (g) | 0.13 (0-1.27) | 0.19 (0.03-0.34) | NS |
| Cholesterol (mg) | 65.5 (30-130) | 324 (259-461) | <.0001 |
| C4:0 (g) | 0.00 (0-0.11) | 0.00 (0-0) | NS |
| C6:0 (g) | 0.00 (0-0.07) | 0.00 (0-0) | NS |
| C8:0 (g) | 0.00 (0-0.04) | 0.00 (0-0.02) | NS |
| C10:0 (g) | 0.01 (0-0.09) | 0.02 (0-0.05) | NS |
| C12:0 (g) | 0.04 (0.01-0.21) | 0.04 (0.01-0.13) | NS |
| C13:0 (g) | 0.00 (0-0) | 0.00 (0-0) | - |
| C14:0 (g) | 0.36 (0.06-0.94) | 0.27 (0.18-0.45) | NS |
| C15:0 (g) | 0.00 (0-0) | 0.02 (0.01-0.02) | <.001 |
| C16:0 (g) | 5.10 (3.04 - 8.57) | 4.81 (3.68-7.16) | NS |
| C17:0 (g) | 0.00 (0-0.01) | 0.01 (0-0.01) | NS |
| C18:0 (g) | 2.25 (1.36-3.23) | 2.25 (1.9-3.72) | NS |
| C20:0 (g) | 0.09 (0.02-0.20) | 0.05 (0.02-0.1) | NS |
| C21:0 (g) | 0.00 (0-0) | 0.00 (0-0) | - |
| C22:0 (g) | 0.05 (0-0.25) | 0.04 (0.02-0.15) | NS |
| C23:0 (g) | 0.00 (0-0) | 0.00 (0-0) | - |
| C24:0 (g) | 0.02 (0-0.10) | 0.04 (0.02-0.10) | NS |
| C10:1 (g) | 0.00 (0-0) | 0.00 (0-0) | - |
| C12:1 (g) | 0.00 (0-0) | 0.00 (0-0) | - |
| C14:1 (g) | 0.00 (0-0.04) | 0.01 (0-0.02) | NS |
| C15:1 (g) | 0.00 (0-0) | 0.00 (0-0) | - |
| C16:1 (g) | 0.33 (0.19-0.53) | 0.60 (0.48-0.97) | 0.01 |
| C17:1 (g) | 0.00 (0-0) | 0.00 (0-0) | NS |
| C18:1 (g) | 10.49 (5.57-18.4) | 10.09 (6.54-14.5) | NS |
| C20:1 (g) | 0.10 (0.03-0.25) | 0.04 (0.02-0.11) | NS |
| C22:1 (g) | 0.04 (0-0.21) | 0.00 (0-0.03) | NS |
| C23:1 (g) | 0.00 (0-0) | 0.00 (0-0) | - |
| C24:1 (g) | 0.00 (0-0) | 0.00 (0-0) | - |
| C18:2 (g) | 5.71 (1.42-35.7) | 5.54 (4.09-12.3) | NS |
| C18:3 (g) | 0.26 (0.11-0.99) | 0.21 (0.14-0.4) | NS |
| C18:4 (g) | 0.00 (0-0.005) | 0.00 (0-0) | NS |
| C20:2 (g) | 0.00 (0-0.01) | 0.04 (0.03-0.06) | 0.001 |
| C20:4 (g) | 0.04 (0-0.07) | 0.07 (0.04-0.1) | NS |
| C20:5 (g) | 0.02 (0-0.04) | 0.03 (0.02-0.04) | NS |
| C22:2 (g) | 0.00 (0-0.003) | 0.01 (0-0.01) | 0.03 |
| C22:3 (g) | 0.00 (0-0) | 0.00 (0-0) | - |
| C22:4 (g) | 0.00 (0-0) | 0.00 (0-0) | - |
| C22:5 (g) | 0.00 (0-0.01) | 0.00 (0-0.01) | NS |
| C22:6 (g) | 0.03 (0-0.07) | 0.07 (0.04-0.08) | NS |
| C24:6 (g) | 0.00 (0-0) | 0.00 (0-0) | - |
| C20:3 (g) | 0.00 (0-0.01) | 0.00 (0-0.01) | NS |
| **Amino acids – Absolute intakes** |  |  |  |
| Isoleucine (g) | 1.46 (0.74-2.10) | 1.41 (1.16-2.16) | NS |
| Leucine (g) | 2.76 (1.38-4.08) | 2.78 (2.14-3.88) | NS |
| Lysine (g) | 1.72 (0.98-2.90) | 2.46 (1.55-3.195) | NS |
| Methionine (g) | 0.83 (0.41-0.96) | 0.92 (0.65-1.19) | NS |
| Phenylalanine (g) | 1.59 (0.79-2.39) | 1.59 (1.21-2.36) | NS |
| Threonine (g) | 1.31 (0.70-1.92) | 1.43 (1.07-1.98) | NS |
| Tryptophan (g) | 0.46 (0.27-0.63) | 0.54 (0.38-0.71) | NS |
| Valine (g) | 1.72 (0.85-2.47) | 1.68 (1.36-2.49) | NS |
| Arginine (g) | 1.69 (1.13-2.83) | 2.23 (1.53-3.27) | NS |
| Histidine (g) | 0.88 (0.41-1.24) | 0.93 (0.73-1.29) | NS |
| Cystine (g) | 0.33 (0.20-0.48) | 0.64 (0.53-0.81) | <.001 |
| Tyrosine (g) | 0.69 (0.29-1.02) | 1.06 (0.73-1.40) | 0.03 |
| Alanine (g) | 0.85 (0.35-1.60) | 1.59 (1.11-2.23) | 0.02 |
| Aspartic acid (g) | 1.45 (0.80-2.69) | 2.90 (1.78-3.53) | 0.04 |
| Glutamic acid (g) | 4.76 (3.04-5.91) | 5.50 (3.31-8.31) | NS |
| Glycine (g) | 0.73 (0.36-1.44) | 1.33 (0.93-2.00) | 0.03 |
| Proline (g) | 1.43 (0.89-2.01) | 1.74 (1.09-2.58) | NS |
| Serine (g) | 0.98 (0.46-1.59) | 1.65 (1.33-2.02) | 0.01 |
| Hydroxyproline (g) | 0.02 (0-0.18) | 0.09 (0-0.17) | NS |
| Alcohol (g) | 0.00 (0-0) | 0.00 (0-0) | - |
| Phytate (mg) | 155 (6-192) | 192 (99.5-225) | NS |
| Malic acid (mg) | 38.0 (0-879) | 305 (14.5-612.5) | NS |
| Citric acid (mg) | 276.50 (0-1275) | 221 (77-508) | NS |
| Tartaric acid (mg) | 0.00 (0-0) | 0.00 (0-0) | NS |
| Oxalic acid (mg) | 6.00 (0-384) | 11.0 (0-37.5) | NS |
| Caffeine (mg) | 0.00 (0-0) | 0.00 (0-0) | - |
| Tannins (mg) | 0.00 (0-0) | 0.00 (0-0) | - |
| **Energy – %** |  |  |  |
| % Energy - Protein | 0.13 (0.08-0.18) | 0.15 (0.13-0.18) | NS |
| % Energy - Fat | 0.20 (0.13-0.40) | 0.25 (0.21-0.31) | NS |
| % Energy - Saturated SFA | 0.06 (0.04-0.07) | 0.06 (0.05-0.09) | NS |
| % Energy - Mono-unsaturated FA | 0.07 (0.05-0.12) | 0.09 (0.08-0.11) | NS |
| % Energy – Polyunsaturated FA | 0.04 (0.02-0.16) | 0.06 (0.04-0.09) | NS |
| % Energy - Carbohydrate | 0.64 (0.52-0.70) | 0.59 (0.51-0.65) | NS |

Data from mother-infant dyads that attended follow up are presented as median (IQR) and p values are from univariate analysis (Kruskal-Wallis/Wilcoxon test for non-parametric data; ANOVA for normal data with equal variance). Iodised salt was assumed to be consumed by all mothers, as the majority of salt consumed in South Africa is iodised (Jooste & Zimmermann, 2008).

**Supplementary table S4.** Maternal nutrient intake from one 24-hour dietary recall for mothers who report experiencing food insecure compared to those who do not experience food insecurity.

|  | Do you worry about food runout? |  |  |
| --- | --- | --- | --- |
|  | NO | YES | p value |
|  | n=14 | n=17 |  |
| Dietary diversity score (/9) | 4.50 (4.00, 6.00) | 4.00 (3.00, 5.50) | NS |
| Dietary diversity score <4 (n) | 2 | 5 | NS |
| **Absolute intakes** |  |  |  |
| Macronutrients |  |  |  |
| Moisture (g) | 868 (629, 1029) | 670 (507, 1092) | NS |
| Energy (kJ) | 5601 (4792, 6855) | 5204 (3264, 8103) | NS |
| Nitrogen (g) | 7.15 (5.55, 8.82) | 4.63 (3.93, 6.57) | NS |
| Total protein (g) | 51.4 (43.8, 63.2) | 37.9 (26.3, 53.8) | NS |
| Plant protein (g) | 19.6 (17.1, 26.0) | 22.3 (11.1, 37.5) | NS |
| Animal protein (g) | 29.0 (19.3, 39.0) | 16.3 (3.85, 30.0) | 0.03 |
| Total fat (g) | 36.5 (30.0, 51.8) | 25.8 (20.6, 65.1) | NS |
| Carbohydrates, avail. (g) | 162 (159, 200) | 161 (97.0, 277) | NS |
| Starch (g) | 5.6 (0.18, 10.3) | 0.00 (0.00, 8.1) | NS |
| Glucose (g) | 3.95 (2.40, 5.00) | 1.80 (0.00, 3.55) | 0.04 |
| Fructose (g) | 6.50 (0.90, 7.45) | 0.70 (0.00, 6.30) | NS |
| Sucrose (g) | 17.7 (12.0, 32.9) | 12.0 (1.70, 23.6) | NS |
| Maltose (g)<sup>1</sup> |  | 0.1 |  |
| Galactose<sup>2</sup> |  |  |  |
| Lactose (g) | 0.00 (0.00, 2.10) | 0.00 (0.00, 0.05) | NS |
| Total sugars (g) | 30.3 (23.4, 41.7) | 15.8 (8.05, 34.5) | NS |
| Added sugar (g) | 25.6 (12.0, 41.0) | 8.00 (0.50, 18.8) | 0.01 |
| Total dietary fibre (g) | 13.4 (11.0, 16.7) | 12.2 (6.70, 21.1) | NS |
| Insoluble dietary fibre (g) | 2.75 (1.33, 3.60) | 1.40 (0.00, 3.45) | NS |
| Soluble dietary fibre (g) | 1.85 (1.05, 2.70) | 0.80 (0.00, 3.40) | NS |
| Ash (g) | 6.05 (4.65, 6.53) | 9.30 (4.30, 14.8) | NS |
| Non-starch polysaccharides (g) | 4.20 (2.13, 5.98) | 2.10 (0.00, 6.45) | NS |
| Insoluble NSP (g) | 2.25 (1.15, 3.03) | 1.40 (0.00, 3.05) | NS |
| Soluble NSP (g) | 1.85 (1.05, 2.70) | 0.80 (0.00, 3.40) | NS |
| Lignin (g) | 0.20 (0.10, 0.30) | 0.10 (0.00, 0.30) | NS |
| Amino acids |  |  |  |
| Isoleucine (g) | 1.78 (1.34, 2.26) | 1.24 (0.80, 1.83) | NS |
| Leucine (g) | 3.48 (2.68, 4.06) | 2.18 (1.51, 3.34) | NS |
| lysine (g) | 2.59 (1.84, 3.67) | 1.52 (1.04, 2.65) | NS |
| Methionine (g) | 0.96 (0.88, 1.17) | 0.65 (0.47, 0.91) | 0.01 |
| Phenylalanine (g) | 2.01 (1.53, 2.35) | 1.32 (0.90, 2.11) | NS |
| Threonine (g) | 1.73 (1.33, 2.10) | 1.08 (0.76, 1.71) | NS |
| Tryptophan (g) | 0.61 (0.53, 0.71) | 0.38 (0.30, 0.57) | 0.02 |
| Valine (g) | 212 (1.67, 2.45) | 1.38 (0.98, 2.23) | NS |
| Arginine (g) | 2.53 (1.86, 3.36) | 1.48 (1.18, 2.77) | NS |
| Histidine (g) | 1.14 (0.85, 1.37) | 0.73 (0.44, 1.27) | 0.05 |
| Cystine (g) | 0.62 (0.53, 0.87) | 0.48 (0.31, 0.66) | 0.02 |
| Tyrosine (g) | 1.12 (0.87, 1.44) | 0.79 (0.46, 1.06) | NS |
| Alanine (g) | 1.74 (1.28, 2.28) | 1.16 (0.67, 1.65) | 0.04 |
| Aspartic acid (g) | 3.01 (2.24, 3.71) | 1.87 (1.12, 2.92) | NS |
| Glutamic acid (g) | 6.24 (4.72, 8.31) | 4.43 (1.96, 5.83) | NS |
| Glycine (g) | 1.47 (1.03, 2.10) | 1.02 (0.52, 1.47) | NS |
| Proline (g) | 2.00 (1.47, 2.63) | 1.36 (0.56, 1.87) | 0.05 |
| Serine (g) | 1.65 (1.50, 2.14) | 1.20 (0.86, 1.67) | 0.03 |
| Hydroxyproline (g) | 0.10 (0.06, 0.22) | 0.00 (0.00, 0.11) | NS |
| Fatty acids and cholesterol |  |  |  |
| Saturated FA (g) | 9.55 (8.56, 15.2) | 7.88 (5.74, 11.4) | NS |
| Mono-unsaturated FA (g) | 12.5 (10.5, 15.8) | 10.9 (7.19, 21.8) | NS |
| Polyunsaturated FA (g) | 8.70 (6.26, 15.3) | 5.46 (3.37, 20.2) | NS |
| Single trans FA (g)<sup>2</sup> |  |  |  |
| Double trans FA (g)<sup>2</sup> |  |  |  |
| Total trans FA (g) | 0.20 (0.16, 0.82) | 0.06 (0.00, 0.35) | NS |
| Cholesterol (mg) | 291 (236, 544) | 117 (33.0, 420) | 0.04 |
| **Median intake (%) of TULs by mothers** |  |  |  |
| Minerals |  |  |  |
| Calcium (Ca) | 5.88 (4.56, 9.55) | 4.52 (2.76, 10.5) | NS |
| Iron (Fe) | 25.6 (21.7, 27.1) | 18.2 (12.8, 31.6) | NS |
| Magnesium (Mg) | 52.1 (44.6, 71.4) | 57.1 (30.0, 97.0) | NS |
| Phosphorus (P) | 16.7 (12.9, 19.9) | 12.1 (9.14, 18.9) | NS |
| Sodium (Na) | 67.5 (48.0, 91.7) | 127 (42.5, 210) | NS |
| Zinc (Zn) | 22.3 (20.1, 29.6) | 19.6 (14.7, 27.1) | NS |
| Copper (Cu) | 8.15 (6.35, 10.6) | 6.90 (3.95, 12.1) | NS |
| Selenium (Se) | 7.56 (5.41, 11.6) | 6.18 (1.86, 9.73) | NS |
| Manganese (Mn) | 12.8 (8.46, 18.2) | 12.7 (5.59, 18.9) | NS |
| Iodine (I) | 7.82 (3.73, 12.8) | 13.6 (4.59, 19.1) | NS |
| Vitamins |  |  |  |
| Vitamin A | 14.2 (11.2, 18.8) | 14.1 (8.57, 26.9) | NS |
| Niacin | 47.4 (42.0, 55.6) | 40.0 (24.4, 60.6) | NS |
| Vitamin B6 | 3.20 (2.74, 3.68) | 2.14 (1.22, 4.17) | NS |
| Folate | 27.8 (21.3, 29.6) | 24.1 (19.8, 48.6) | NS |
| Vitamin C | 1.28 (0.90, 2.45) | 0.60 (0.08, 6.38) | NS |
| Vitamin D | 4.73 (4.15, 8.76) | 1.39 (0.39, 7.86) | NS |
| Vitamin E | 0.73 (0.49, 1.21) | 0.68 (0.41, 1.30) | NS |
| **Do you experience food runout?** |  |  |  |
|  | **NO** | **YES** | **p value** |
|  | n=16 | n=15 |  |
| **Dietary diversity score (/9)** | 5.00 (4.00, 6.00) | 3.00 (4.00, 5.00) | NS |
| **Dietary diversity score <4 (n)** | 2 | 5 | NS |
| **Absolute intakes** |  |  |  |
| Macronutrients |  |  |  |
| Moisture (g) | 868 (620, 1030) | 670 (409, 1011) | NS |
| Energy (kJ) | 5601 (4659, 7004) | 5204 (3247, 8339) | NS |
| Nitrogen (g) | 7.15 (5.68, 9.29) | 4.31 (3.87, 6.10) | 0.02 |
| Total protein (g) | 51.4 (42.5, 68.7) | 32.9 (26.1, 44.7) | 0.03 |
| Plant protein (g) | 19.6 (15.8, 27.3) | 22.3 (11.5, 34.9) | NS |
| Animal protein (g) | 29.0 (19.7, 38.0) | 15.4 (3.30, 21.4) | 0.01 |
| Total fat (g) | 35.0 (29.9, 45.8) | 25.8 (19.6, 69.0) | NS |
| Carbohydrates, avail. (g) | 162 (159, 234) | 161 (93.8, 280) | NS |
| Starch (g) | 5.60 (0.13, 10.1) | 0.00 (0.00, 5.40) | NS |
| Glucose (g) | 3.95 (2.75, 5.60) | 1.20 (0.00, 3.40) | 0.01 |
| Fructose (g) | 6.50 (1.20, 8.15) | 0.40 (0.00, 6.10) | 0.01 |
| Sucrose (g) | 19.1 (12.4, 30.1) | 8.40 (1.00, 17.9) | 0.03 |
| Maltose (g)<sup>1</sup> |  | 0.1 |  |
| Galactose<sup>2</sup> |  |  |  |
| Lactose (g) | 0.00 (0.00, 1.60) | 0.00 (0.00, 0.10) | NS |
| Total sugars (g) | 31.2 (24.3, 42.5) | 13.1 (7.00, 30.9) | 0.02 |
| Added sugar (g) | 17.6 (12.0, 40.5) | 8.00 (0.30, 21.0) | 0.04 |
| Total dietary fibre (g) | 13.4 (10.9, 17.2) | 12.2 (6.20, 18.4) | NS |
| Insoluble dietary fibre (g) | 3.15 (1.68, 4.08) | 1.20 (0.00, 3.20) | 0.04 |
| Soluble dietary fibre (g) | 2.20 (1.30, 2.70) | 0.70 (0.00, 2.90) | NS |
| Ash (g) | 6.05 (4.55, 6.58) | 10.5 (4.20, 14.8) | NS |
| Non-starch polysaccharides (g) | 4.85 (2.68, 6.33) | 1.80 (0.00, 5.90) | NS |
| Insoluble NSP (g) | 2.60 (1.43, 3.50) | 1.10 (0.00, 2.80) | NS |
| Soluble NSP (g) | 2.20 (1.30, 2.70) | 0.70 (0.00, 2.90) | NS |
| Lignin (g) | 0.25 (0.13, 0.38) | 0.00 (0.00, 0.20) | 0.01 |
| Amino acids |  |  |  |
| Isoleucine (g) | 1.78 (1.38, 2.43) | 1.08 (0.79, 1.47) | 0.05 |
| Leucine (g) | 3.48 (2.77, 4.47) | 1.95 (1.47, 2.75) | 0.03 |
| lysine (g) | 2.59 (1.99, 3.80) | 1.23 (1.03, 2.57) | 0.03 |
| Methionine (g) | 0.96 (0.88, 1.13) | 0.61 (0.45, 0.85) | 0.002 |
| Phenylalanine (g) | 2.01 (1.55, 2.51) | 1.22 (0.80, 1.69) | 0.04 |
| Threonine (g) | 1.73 (1.38, 2.22) | 0.95 (0.74, 1.43) | 0.02 |
| Tryptophan (g) | 0.61 (0.53, 0.76) | 0.34 (0.30, 0.51) | 0.003 |
| Valine (g) | 2.12 (1.68, 2.56) | 1.24 (0.87, 1.75) | 0.04 |
| Arginine (g) | 2.53 (1.96, 3.57) | 1.48 (1.17, 2.12) | 0.03 |
| Histidine (g) | 1.14 (0.89, 1.51) | 0.64 (0.42, 1.26) | 0.01 |
| Cystine (g) | 0.62 (0.52, 0.81) | 0.48 (0.30, 0.62) | 0.02 |
| Tyrosine (g) | 1.14 (0.94, 1.42) | 0.75 (0.46, 0.90) | 0.03 |
| Alanine (g) | 1.75 (1.37, 2.29) | 0.92 (0.66, 1.46) | 0.006 |
| Aspartic acid (g) | 3.01 (2.46, 3.74) | 1.47 (1.04, 2.90) | 0.04 |
| Glutamic acid (g) | 5.85 (5.25, 8.12) | 4.33 (1.70, 5.78) | NS |
| Glycine (g) | 1.52 (1.13, 2.14) | 0.84 (0.42, 1.30) | 0.009 |
| Proline (g) | 1.85 (1.58, 2.59) | 1.28 (0.51, 1.92) | 0.03 |
| Serine (g) | 1.68 (1.52, 2.02) | 1.09 (0.85, 1.48) | 0.01 |
| Hydroxyproline (g) | 0.11 (0.06, 0.26) | 0.00 (0.00, 0.07) | 0.005 |
| Fatty acids and cholesterol |  |  |  |
| Saturated FA (g) | 9.68 (8.88, 14.7) | 6.80 (5.69, 12.9) | 0.03 |
| Mono-unsaturated FA (g) | 11.7 (10.4, 15.0) | 12.0 (6.86, 25.7) | NS |
| Polyunsaturated FA (g) | 7.67 (5.87, 13.9) | 5.47 (3.88, 22.7) | NS |
| Single trans FA (g)<sup>2</sup> |  |  |  |
| Double trans FA (g)<sup>2</sup> |  |  |  |
| Total trans FA (g) | 0.21 (0.16, 0.75) | 0.04 (0.00, 0.20) | 0.03 |
| Cholesterol (mg) | 291 (191, 524) | 112 (26.0, 416) | 0.03 |
| **Median intake (%) of TULs by mothers** |  |  |  |
| **Minerals** |  |  |  |
| Calcium (Ca) | 5.88 (4.56, 9.90) | 4.52 (2.72, 9.56) | NS |
| Iron (Fe) | 25.6 (21.0, 27.4) | 18.2 (12.0, 30.2) | NS |
| Magnesium (Mg) | 52.1 (43.1, 74.6) | 57.1 (30.3, 95.1) | NS |
| Phosphorus (P) | 16.7 (12.8, 20.7) | 12.1 (8.65, 18.5) | NS |
| Sodium (Na) | 61.9 (47.6, 84.9) | 132 (51.2, 221) | NS |
| Zinc (Zn) | 23.3 (20.2, 30.3) | 17.7 (14.4, 24.2) | NS |
| Copper (Cu) | 8.15 (6.43, 11.7) | 6.90 (3.90, 11.0) | NS |
| Selenium (Se) | 7.63 (5.91, 11.2) | 5.58 (1.80, 7.78) | NS |
| Manganese (Mn) | 12.8 (8.77, 19.3) | 12.7 (5.36, 16.7) | NS |
| Iodine (I) | 7.67 (3.73, 11.2) | 14.0 (5.09, 19.6) | NS |
| **Vitamins** |  |  |  |
| Vitamin A | 14.2 (10.5, 20.9) | 14.1 (8.54, 25.2) | NS |
| Niacin | 47.4 (41.4, 55.9) | 40.0 (23.4, 55.7) | NS |
| Vitamin B6 | 3.20 (2.72, 3.89) | 1.89 (1.16, 3.80) | NS |
| Folate | 27.8 (20.0, 30.1) | 24.1 (20.5, 47.6) | NS |
| Vitamin C | 1.34 (0.99, 2.55) | 0.55 (0.00, 6.25) | NS |
| Vitamin D | 4.73 (4.14, 8.60) | 1.39 (0.05, 7.86) | NS |
| Vitamin E | 0.73 (0.49, 1.16) | 0.68 (0.39, 1.46) | NS |
| **Are you able to afford balanced meals?** |  |  |  |
|  | **YES** | **NO** | **p value** |
|  | n=9 | n=22 |  |
| **Dietary diversity score (/9)** | 4.00 (3.50, 5.50) | 4.00 (3.75, 6.00) | NS |
| **Dietary diversity score <4 (n)** | 2 | 5 | NS |
| **Absolute intakes** |  |  |  |
| **Macronutrients** |  |  |  |
| Moisture (g) | 789 (614, 1037) | 806 (564, 1016) | NS |
| Energy (kJ) | 5691 (4519, 7046) | 5358 (3538, 7505) | NS |
| Nitrogen (g) | 6.94 (5.00, 9.53) | 5.52 (4.03, 7.37) | NS |
| Total protein (g) | 53.3 (41.5, 67.7) | 40.9 (27.5, 54.6) | NS |
| Plant protein (g) | 19.9 (17.0, 33.1) | 20.6 (11.6, 34.0) | NS |
| Animal protein (g) | 28.5 (19.0, 38.5) | 18.9 (9.65, 33.2) | NS |
| Total fat (g) | 37.5 (25.1, 57.2) | 31.0 (21.9, 63.1) | NS |
| Carbohydrates, avail. (g) | 165 (158, 229) | 160 (106, 259) | NS |
| Starch (g) | 5.60 (0.05, 6.80) | 0.20 (0.00, 12.5) | NS |
| Glucose (g) | 3.80 (2.50, 5.25) | 2.55 (0.30, 4.55) | NS |
| Fructose (g) | 6.50 (1.90, 7.75) | 1.10 (0.00, 6.65) | NS |
| Sucrose (g) | 20.4 (16.3, 28.1) | 12.0 (3.98, 23.9) | NS |
| Maltose (g)<sup>1</sup> |  | 0.1 |  |
| Galactose<sup>2</sup> |  |  |  |
| Lactose (g) | 0.00 (0.00, 2.40) | 0.00 (0.00, 0.15) | NS |
| Total sugars (g) | 32.0 (29.6, 39.5) | 20.8 (9.10, 37.1) | NS |
| Added sugar (g) | 13.4 (10.5, 25.6) | 12.0 (3.70, 39.5) | NS |
| Total dietary fibre (g) | 14.5 (11.6, 16.3) | 12.3 (8.10, 17.9) | NS |
| Insoluble dietary fibre (g) | 3.10 (1.35, 3.90) | 1.75 (0.00, 3.33) | NS |
| Soluble dietary fibre (g) | 2.20 (0.85, 2.75) | 1.30 (0.00, 2.75) | NS |
| Ash (g) | 6.10 (4.25, 6.55) | 6.15 (4.63, 14.6) | NS |
| Non-starch polysaccharides (g) | 4.90 (2.00, 6.25) | 2.75 (0.00, 6.05) | NS |
| Insoluble NSP (g) | 2.50 (1.05, 3.30) | 1.45 (0.00, 2.98) | NS |
| Soluble NSP (g) | 2.20 (0.85, 2.75) | 0.00 (1.30, 2.75) | NS |
| Lignin (g) | 0.30 (0.20, 0.40) | 0.10 (0.00, 0.23) | NS |
| Amino acids |  |  |  |
| Isoleucine (g) | 1.77 (1.26, 2.55) | 1.50 (0.89, 1.95) | NS |
| Leucine (g) | 3.36 (2.60, 2.46) | 2.47 (1.58, 3.71) | NS |
| lysine (g) | 2.71 (1.81, 4.05) | 1.74 (1.06, 2.65) | NS |
| Methionine (g) | 1.06 (0.93, 2.44) | 1.01 (0.78, 1.64) | NS |
| Phenylalanine (g) | 1.96 (1.42, 2.43) | 1.51 (1.00, 2.22) | NS |
| Threonine (g) | 1.69 (1.26, 2.23) | 1.25 (0.79, 1.81) | NS |
| Tryptophan (g) | 0.59 (0.48, 0.77) | 0.42 (0.31, 0.63) | NS |
| Valine (g) | 2.01 (1.53, 2.58) | 1.57 (1.12, 2.38) | NS |
| Arginine (g) | 2.44 (1.79, 3.49) | 1.80 (1.28, 2.68) | NS |
| Histidine (g) | 1.16 (0.82, 1.51) | 0.76 (0.50, 1.20) | NS |
| Cystine (g) | 0.60 (0.48, 0.78) | 0.52 (0.34, 0.72) | NS |
| Tyrosine (g) | 1.09 (0.80, 1.40) | 0.85 (0.49, 1.10) | NS |
| Alanine (g) | 1.76 (1.20, 2.32) | 1.24 (0.73, 1.73) | NS |
| Aspartic acid (g) | 2.93 (2.06, 3.90) | 1.87 (1.30, 3.02) | NS |
| Glutamic acid (g) | 5.71 (4.26, 7.54) | 4.58 (2.39, 6.46) | NS |
| Glycine (g) | 1.49 (1.06, 2.13) | 1.04 (0.63, 1.48) | NS |
| Proline (g) | 1.81 (1.36, 2.35) | 1.38 (0.78, 2.36) | NS |
| Serine (g) | 1.65 (1.29, 2.02) | 1.43 (0.91, 1.71) | NS |
| Hydroxyproline (g) | 0.11 (0.07, 0.24) | 0.00 (0.00, 0.11) | 0.04 |
| Fatty acids and cholesterol |  |  |  |
| Saturated FA (g) | 9.77 (8.33, 19.5) | 7.96 (5.98, 12.1) | NS |
| Mono-unsaturated FA (g) | 11.8 (8.26, 16.4) | 11.5 (7.6, 21.8) | NS |
| Polyunsaturated FA (g) | 9.40 (4.85, 15.5) | 6.34 (4.49, 18.4) | NS |
| Single trans FA (g)<sup>2</sup> |  |  |  |
| Double trans FA (g)<sup>2</sup> |  |  |  |
| Total trans FA (g) | 0.73 (0.16, 1.00) | 0.06 (0.00, 0.21) | 0.03 |
| Cholesterol (mg) | 269 (213, 326) | 261 (45.5, 446) | NS |
| **Median intake (%) of TULs by mothers** |  |  |  |
| Minerals |  |  |  |
| Calcium (Ca) | 7.04 (4.90, 11.7) | 4.76 (3.01, 8.33) | NS |
| Iron (Fe) | 25.8 (213, 30.0) | 24.2 (14.6, 28.3) | NS |
| Magnesium (Mg) | 54.0 (46.4, 78.7) | 52.1 (30.7, 88.6) | NS |
| Phosphorus (P) | 15.2 (12.9, 21.2) | 14.6 (9.83, 18.9) | NS |
| Sodium (Na) | 65.1 (48.4, 95.0) | 73.6 (45.0, 181) | NS |
| Zinc (Zn) | 20.9 (20.2, 36.8) | 21.0 (15.2, 26.6) | NS |
| Copper (Cu) | 8.60 (6.30, 10.7) | 7.15 (4.45, 11.4) | NS |
| Selenium (Se) | 7.50 (6.03, 8.66) | 6.45 (2.73, 9.89) | NS |
| Manganese (Mn) | 14.7 (8.84, 27.1) | 11.5 (6.52, 16.9) | NS |
| Iodine (I) | 8.18 (3.59, 13.6) | 8.36 (4.84, 18.4) | NS |
| Vitamins |  |  |  |
| Vitamin A | 12.2 (10.2, 16.1) | 14.7 (8.95, 24.9) | NS |
| Niacin | 46.6 (41.7, 54.0) | 42.1 (24.9, 58.1) | NS |
| Vitamin B6 | 3.26 (2.80, 5.09) | 2.61 (1.37, 3.69) | NS |
| Folate | 27.8 (22.6, 29.9) | 25.0 (19.3, 37.1) | NS |
| Vitamin C | 2.40 (0.85, 3.90) | 1.13 (0.23, 3.59) | NS |
| Vitamin D | 4.72 (4.20, 6.22) | 4.12 (0.72, 8.06) | NS |
| Vitamin E | 0.80 (0.51, 1.13) | 0.62 (0.42, 1.50) | NS |

Data from mother-infant dyads that attended follow up are presented as mean ± SD or median (IQR) and p values are from univariate analysis (Kruskal-Wallis/Wilcoxon test for non-parametric data; ANOVA for normal data with equal variance). Dietary diversity score was calculated using nine food groups, only counting each food groups once (refs). Percent intake of TULs were calculated using the Institute of Medicine's TULs for minerals and vitamins for lactating women 14-18, 19-30 or 31-50 years of age<sup>26</sup>. Iodised salt was assumed to be consumed by all mothers, as the majority of salt consumed in South Africa is iodised<sup>64</sup>. TULs = Tolerable upper levels. <sup>1</sup>Only one mother consumed maltose. <sup>2</sup>None consumed by mothers.

**Supplementary figure S1.**
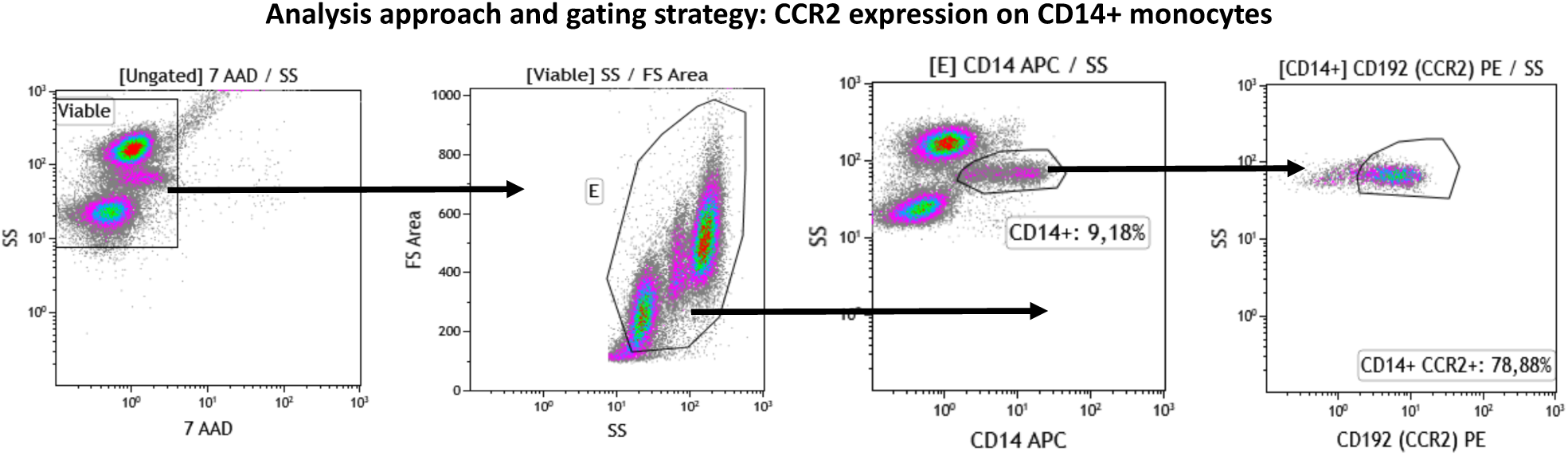
Sequential gating approach for the measurement of CCR2 expression by CD14+ monocytes. The sequential gating approached used was as follows: First, the viable (7-AAD negative; region ‘Viable’’) cells were identified using a 7-AAD vs SS Log density plot. A “Viable” region was created around the 7-AAD negative cells. Gated on the “Viable”cells, a SSLog vs FS plot was used to capture intact cells in the “E” region. CD14+ monocytes were identified (“CD14+” region) using a CD14 vs SS Log density plot that were gated on viable, intact cells (“E” region). CD14+ monocytes that express CCR2 were quantified using a CD192 (CCR2) vs SS Log plot. The proportion of CD14+/CCR2+ cells were captured in the “CD14+ CCR2+” region. The gating strategy followed to quantify CCR2 expression by CD16+ neutrophils was similar to what was described for CD14+ monocytes, but instead of identifying CD14+ monocytes, CD16+ neutrophils were identified (“CD16+” region) using a CD16 vs SS Log density plot that were gated on viable, intact cells (“E” region). CD16+ neutrophils that express CCR2 were quantified using a CD192 (CCR2) vs SS Log plot. The proportion of CD14+/CCR2+ cells was captured in the “CD16+ CCR2+” region.

**Supplementary figure S2.**
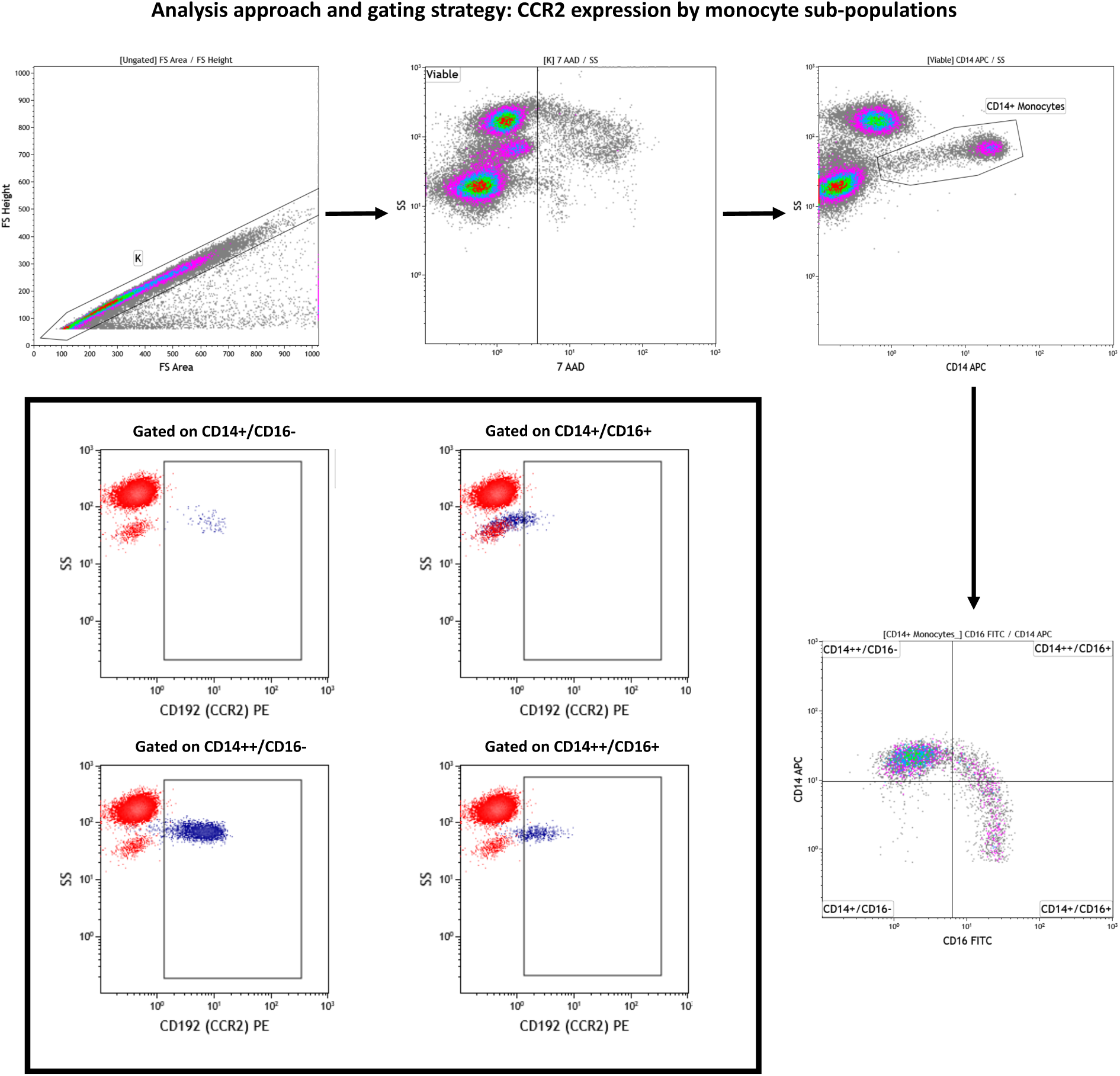
Sequential gating approach for the measurement of CCR2 expression by monocyte sub-populations. Doublets and debris were removed (Region ‘K’) using a FS Area vs FS Height density plot. A 7-AAD vs SS Log density plot, gated on ‘K’ was used to exclude all non-viable cells. Viable cells were captured in region ‘Viable’. Viable CD14+ monocytes were identified (Region ‘CD14+ Monocytes’) using a CD14 APC vs SS Log density plot. Monocyte sub-populations were identified using a CD16 FITC vs CD14 PE density plot gated on viable, CD14+ monocytes. Four monocyte sub-populations were identified: CD14+/CD16-; CD14++/CD16-; CD14+/CD16+; and CD14++/CD16+. The percentage CCR2^+^ monocytes present in each of the respective monocyte sub-populations were identified using CD195 (CCR2) PE vs SS Log two-parameter plots gated on the respective sub-populations. The overlay plots within the black bordered square indicates the strategy used to determine CCR2 expression of the different monocyte subsets. The negative/positive staining boundaries were determined based on the negative expression of CCR2 by CD16^++^/CD14^-^neutrophils (indicated in red in the overlay plots). The CCR2^+^ populations are indicated in blue.

**Supplementary figure S3.**
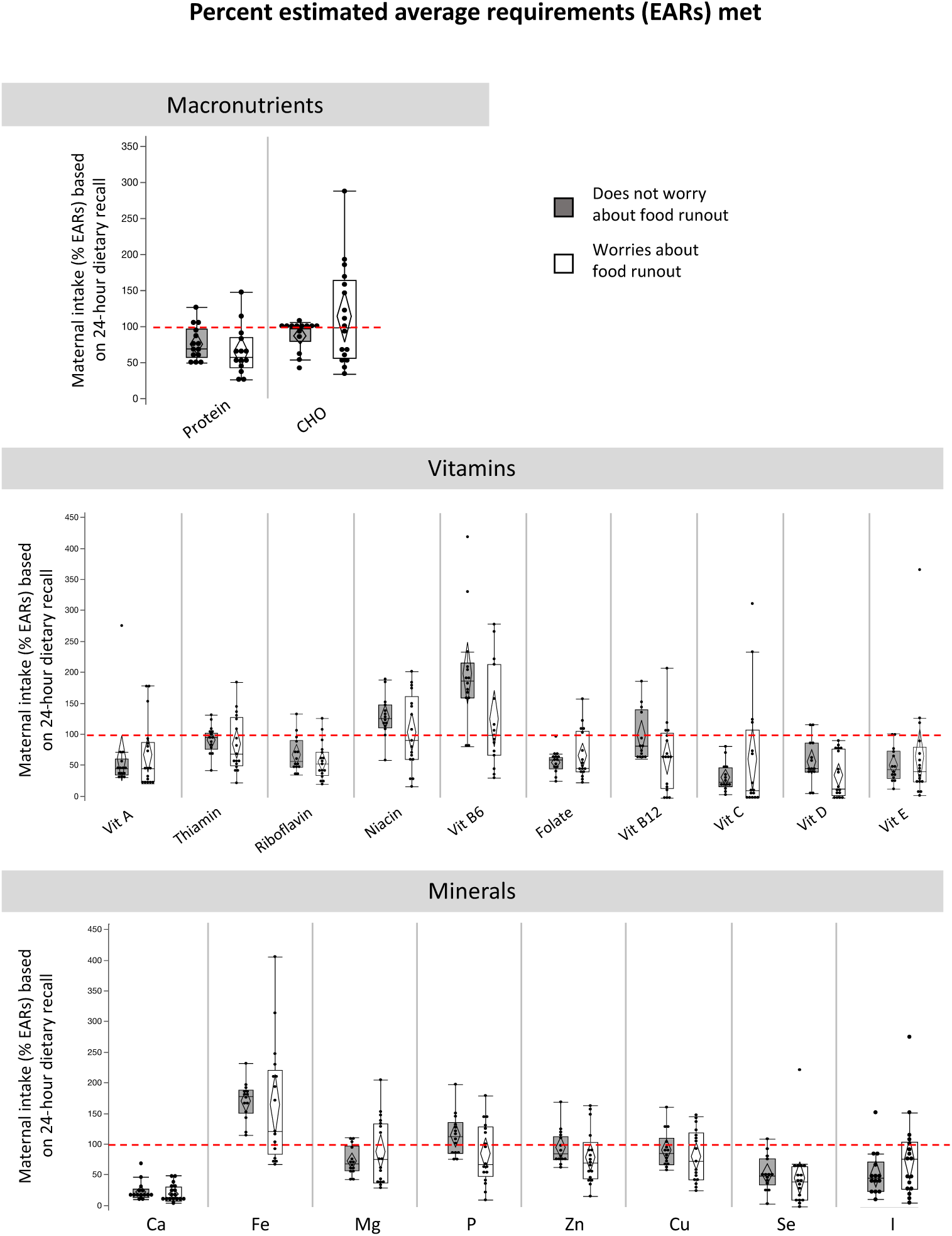
Maternal intake of estimated average requirements for macronutrients, vitamins and minerals for mothers who report worrying about food runout (compared to not worrying). Maternal reports of food insecurity did not associate with intake levels of macronutrients, vitamins or minerals. Many women, irrespective of food security reports, are at risk of inadequate macronutrient, vitamin and mineral intakes. Percent intake of EARs for nutrients were calculated for lactating women 14-18, 19-30 or 31-50 years of age^36^. No EARs are available for total fat. Calculations for EAR for total protein considered maternal weight at time of dietary recall. Data are % intake of EAR reported in maternal dietary recall for macronutrients (quartiles, median lines and 95% confidence diamonds, *p<0.05 [ANOVA for normal distribution/equal variance; Kruskal-Wallis/Wilcoxon test for non-parametric data; or Welch’s test for normal data/unequal variance]). CHO = carbohydrates.

**Supplementary figure S4.**
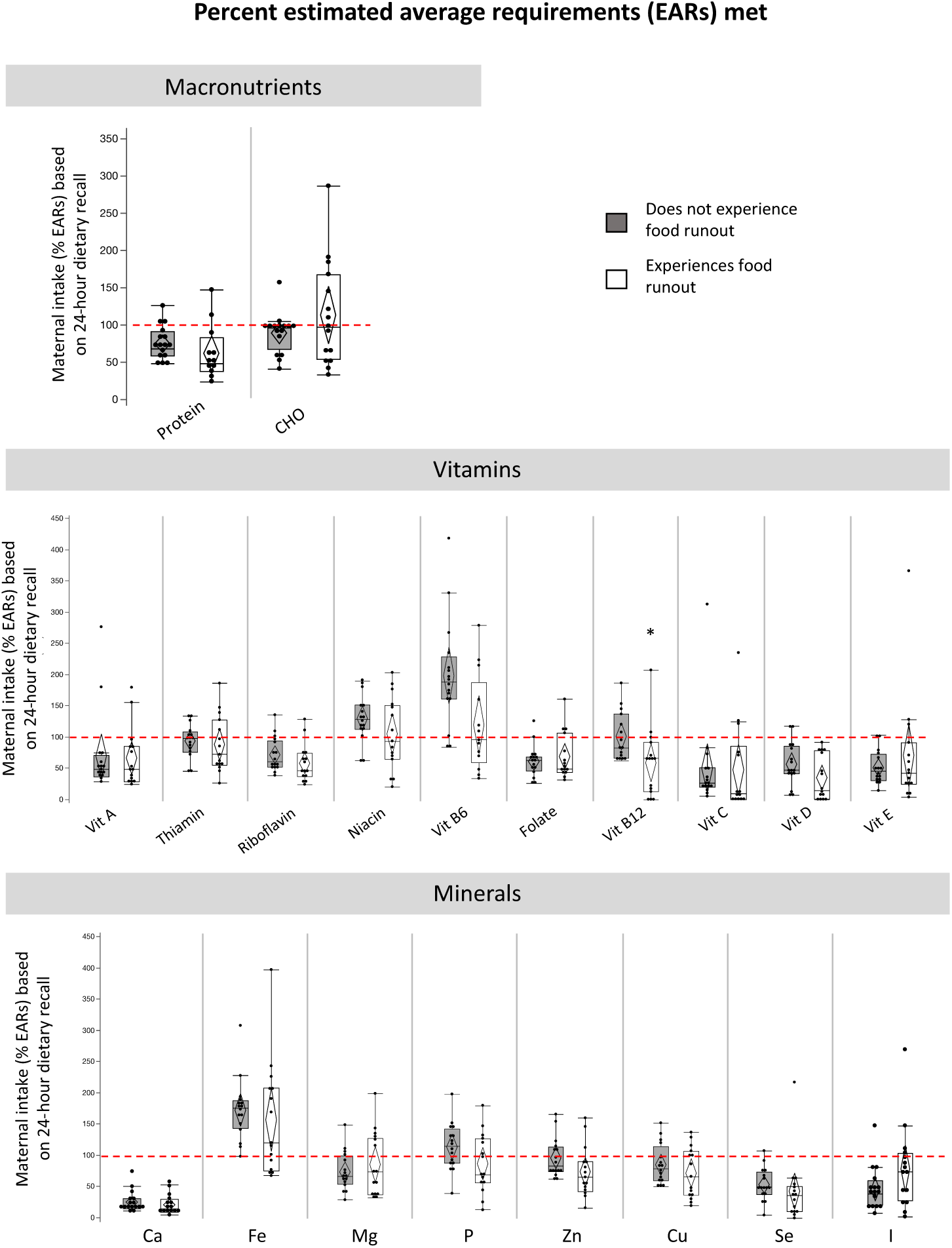
Maternal intake of estimated average requirements for macronutrients, vitamins and minerals for mothers who report experiencing food runout (compared to not). Maternal reports of food insecurity did not associate with intake levels of macronutrients or minerals. Maternal reports of experiencing food runout associated with lower intake of vitamin B12 [B; AdjP=0.20, p=0.01]). Many women, irrespective of food security reports, are at risk of inadequate macronutrient, vitamin and mineral intakes. Percent intake of EARs for nutrients were calculated for lactating women 14-18, 19-30 or 31-50 years of age^36^. No EARs are available for total fat. Calculations for EAR for total protein considered maternal weight at time of dietary recall. Data are % intake of EAR reported in maternal dietary recall for macronutrients (quartiles, median lines and 95% confidence diamonds, *p<0.05 [ANOVA for normal distribution/equal variance; Kruskal-Wallis/Wilcoxon test for non-parametric data; or Welch’s test for normal data/unequal variance]). CHO = carbohydrates.

**Supplementary figure S5.**
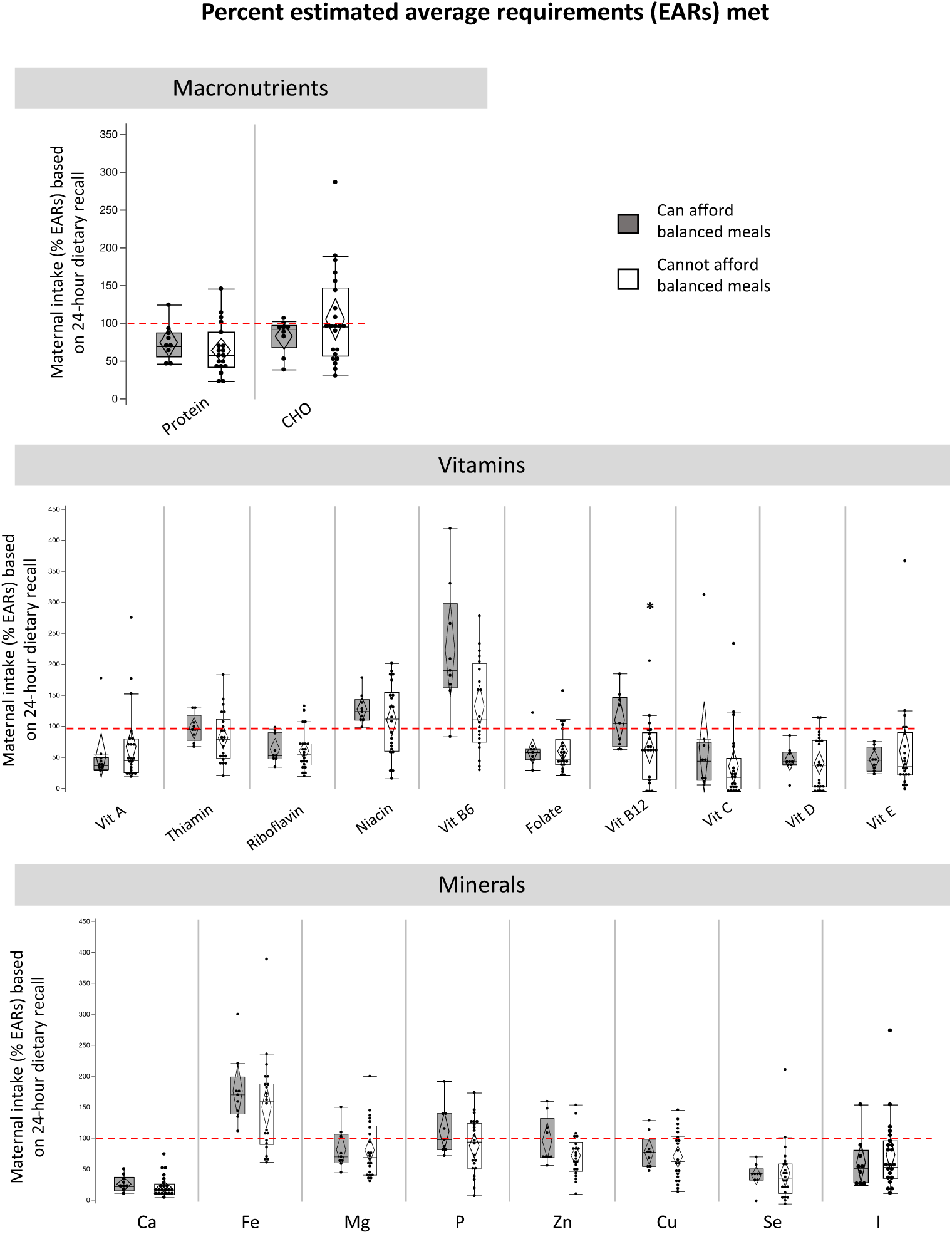
Maternal intake of estimated average requirements for macronutrients, vitamins and minerals for mothers who report inability to afford balanced meals (compared to not). Maternal reports of food insecurity did not associate with intake levels of macronutrients or minerals. Maternal reports of inability to afford balanced meals associated with lower intake of vitamin B12 (C; AdjP=0.05, p=0.04). Many women, irrespective of food security reports, are at risk of inadequate macronutrient, vitamin and mineral intakes. Percent intake of EARs for nutrients were calculated for lactating women 14-18, 19-30 or 31-50 years of age^36^. No EARs are available for total fat. Calculations for EAR for total protein considered maternal weight at time of dietary recall. Data are % intake of EAR reported in maternal dietary recall for macronutrients (quartiles, median lines and 95% confidence diamonds, *p<0.05 [ANOVA for normal distribution/equal variance; Kruskal-Wallis/Wilcoxon test for non-parametric data; or Welch’s test for normal data/unequal variance]). CHO = carbohydrates.

**Supplementary figure S6.**
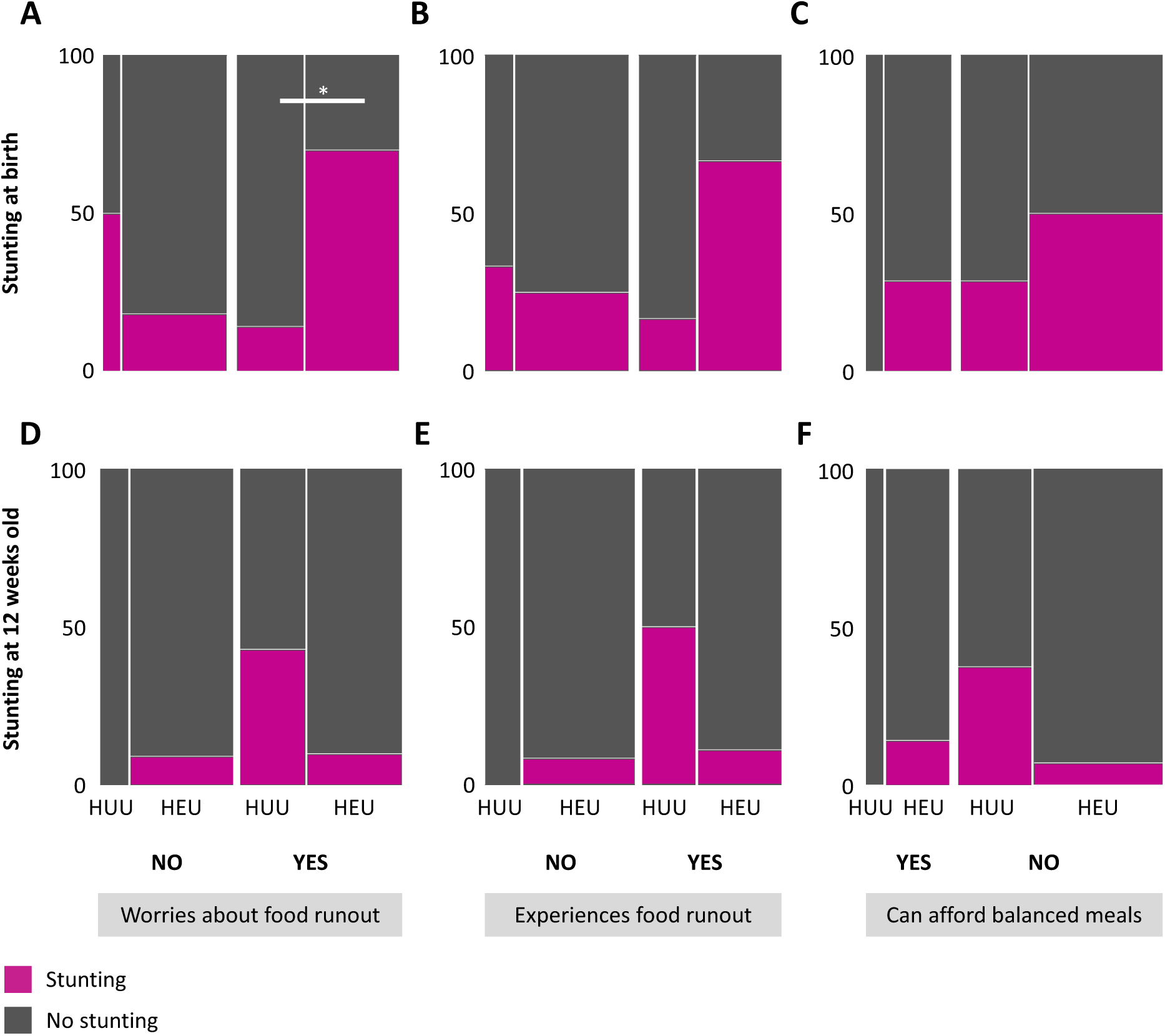
Cooccurrence of maternal HIV and food insecurity may increase risk of stunting at birth. Amongst infants whose mothers report worrying about food runout, risk of stunting at birth is greater for HEU compared to HUU infants (*e*; p=0.04, Fisher’s exact test). The red line represents the proportion of infants who had stunting at birth or 12 weeks PP. Mosaic plots are proportion (%) of HUU or HEU infants who have stunting (<-2 SD length-for-age standardised according to WHO child growth standards^27^) at birth and 12 weeks old. HUU = HIV-unexposed, uninfected infant; HEU = HIV-exposed, uninfected infant.

**Supplementary figure S7.**
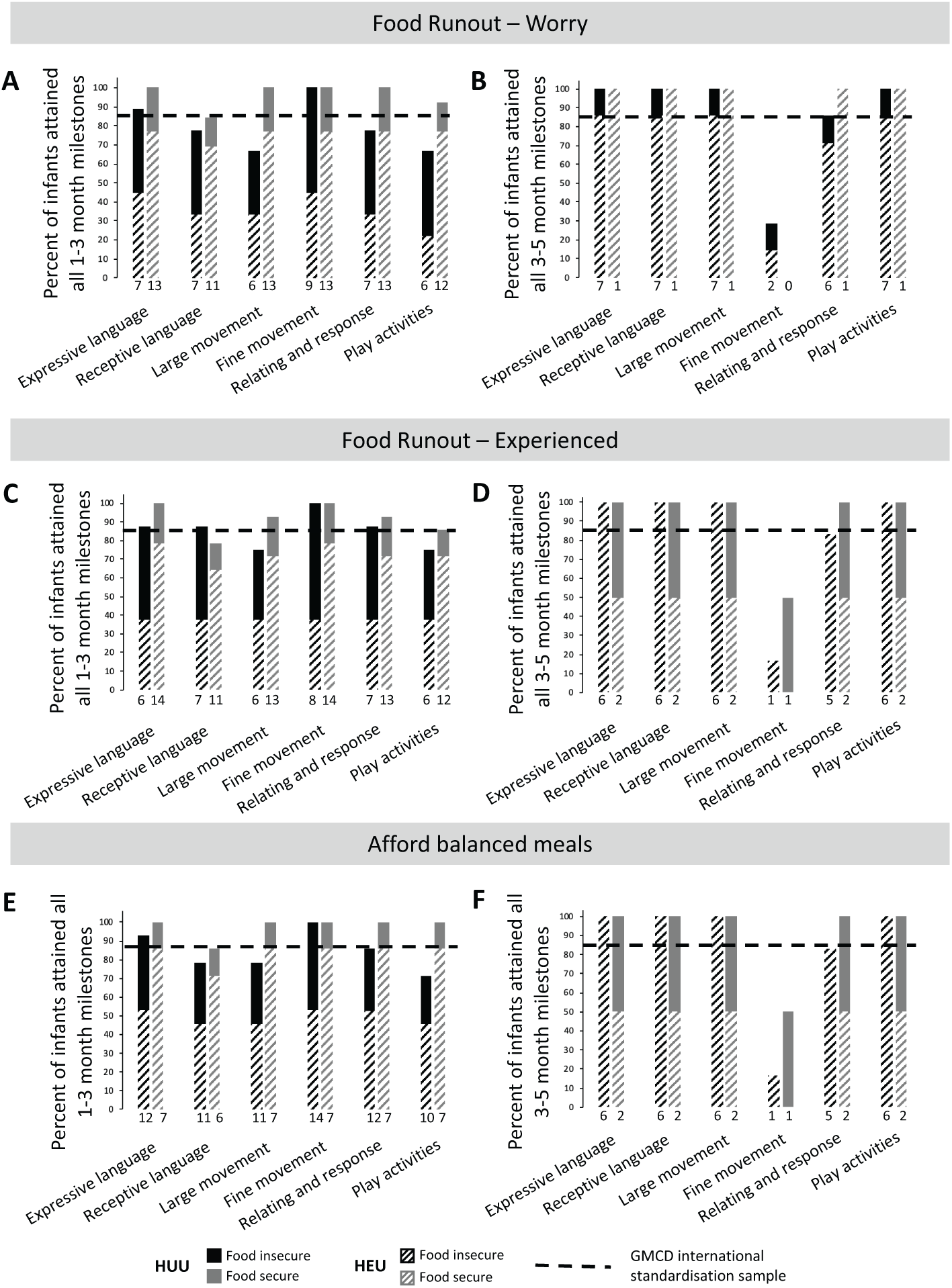
Food insecurity may associate with low attainment of GMCD milestones for HUU and HEU infants. Infants whose mothers reported household food insecurity did not attain 1-3 month GMCD milestones (A, C, E) for receptive language, large movement, relating and response behaviour or play activities, or 3-5 month GMCD milestones (B, D, F) for fine movement or relating and response behaviour in the same proportion as the international standardization sample. Maternal reports of food insecurity did not associate with risk of not attaining all 1-3 month or 3-5 month GMCD milestones (A-F, [p>0.05], Fisher’s exact 2-Tail). Data are proportion (%) of infants who attained all age-appropriate GMCD milestones. The horizontal dotted line represents the GMCD standardised international sample proportion (85%) of infants who attained all milestones in that age category, when they were in that age range. The numbers underneath the bars represent the number of infants attaining all milestones for each milestone. GMCD = Guide for monitoring child development; HUU = HIV-unexposed, uninfected infant; HEU = HIV-exposed, uninfected infant.

## References

1. Gibb, D. M. et al. Pregnancy and infant outcomes among HIV-infected women taking long-term ART with and without tenofovir in the DART trial. PLoS Med 9, e1001217 (2012).

2. UNAIDS. The Gap Report. (2014).

3. Rollins, N. C. et al. Pregnancy Outcomes in HIV-Infected and Uninfected Women in Rural and Urban South Africa. JAIDS J. Acquir. Immune Defic. Syndr. 44, 321–328 (2007).

4. Arab, K., Spence, A. R., Czuzoj-Shulman, N. & Abenhaim, H. A. Pregnancy outcomes in HIV-positive women: a retrospective cohort study. Arch. Gynecol. Obstet. 295, 599–606 (2017).

5. Dadhwal, MD, V. et al. Pregnancy Outcomes in HIV-Infected Women: Experience from a Tertiary Care Center in India. Int. J. MCH AIDS 6, 75 (2017).

6. Faa, G. et al. Fetal programming of neuropsychiatric disorders. Birth Defects Res. Part C Embryo Today Rev. 108, 207–223 (2016).

7. Drotar, D. et al. Neurodevelopmental outcomes of Ugandan infants with HIV infection: An application of growth curve analysis. Heal. Psychol. 18, 114–121 (1999).

8. Boivin, M. J. et al. A preliminary evaluation of the cognitive and motor effects of pediatric HIV infection in Zairian children. Health Psychol. 14, 13–21 (1995).

9. Msellati, P. et al. Neurodevelopmental testing of children born to human immunodeficiency virus type 1 seropositive and seronegative mothers: a prospective cohort study in Kigali, Rwanda. Pediatrics 92, 843–8 (1993).

10. UNAIDS. Global AIDS Monitoring 2018. (2018).

11. South African National AIDS Council (SANAC). Let our actions count: National Strategic Plan on HIV, TB and STIs (2017-2022). (2018).

12. UNAIDS. UNAIDS Country factsheets South Africa 2018. (2018).

13. Volmink, J., Siegfried, N. L., Merwe, L. van der & Brocklehurst, P. Antiretrovirals for reducing the risk of mother-to-child transmission of HIV infection. Cochrane Database Syst. Rev. (2007). doi:10.1002/14651858.CD003510.pub2

14. da Silva, K. M., de Sá, C. dos S. C. & Carvalho, R. Evaluation of motor and cognitive development among infants exposed to HIV. Early Hum. Dev. 105, 7–10 (2017).

15. Springer, P. E. et al. Neurodevelopmental outcome of HIV-exposed but uninfected infants in the Mother and Infants Health Study, Cape Town, South Africa. Trop. Med. Int. Heal. 23, 69–78 (2018).

16. Wu, J. et al. Neurodevelopmental outcomes in young children born to HIV-positive mothers in rural Yunnan, China. Pediatr. Int. (2018). doi:10.1111/ped.13584

17. Funderburg, N. T. et al. Shared monocyte subset phenotypes in HIV- 1 infection and in uninfected subjects with acute coronary syndrome. Blood 120, 4599–608 (2012).

18. Saylor, D. et al. HIV-associated neurocognitive disorder — pathogenesis and prospects for treatment. Nat. Rev. Neurol. 12, 234–248 (2016).

19. Hemkens, L. G. & Bucher, H. C. HIV infection and cardiovascular disease. Eur. Heart J. 35, 1373–1381 (2014).

20. Veenstra, M. et al. CCR2 on Peripheral Blood CD14+CD16+ Monocytes Correlates with Neuronal Damage, HIV-Associated Neurocognitive Disorders, and Peripheral HIV DNA: reseeding of CNS reservoirs? J. Neuroimmune Pharmacol. 14, 120–133 (2019).

21. Uprety, P. et al. Inflammation and Immune Activation in Antiretroviral- Treated Human Immunodeficiency Virus Type 1–Infected African Infants and Rotavirus Vaccine Responses. J. Infect. Dis. 215, 928 (2017).

22. Statistics South Africa. Poverty on the rise in South Africa. (2017).

23. Georgieff, M. K. Nutrition and the developing brain: nutrient priorities and measurement. Am. J. Clin. Nutr. 85, 614S–620S (2007).

24. Barker, D. J. P. & Thornburg, K. L. The Obstetric Origins of Health for a Lifetime. Clin. Obstet. Gynecol. 56, 511–519 (2013).

25. Cusick, S. E. & Georgieff, M. K. The Role of Nutrition in Brain Development: The Golden Opportunity of the ‘First 1000 Days’. J. Pediatr. 175, 16–21 (2016).

26. Western Cape Government, D. of H. Road to Health Card | Western Cape Government. Western Cape Government (2014). Available at: https://www.westerncape.gov.za/general-publication/road-health-card. (Accessed: 19th May 2019)

27. WHO | WHO Anthro (version 3.2.2, January 2011) and macros. WHO (2017).

28. McLennan, J. E., Gilles, F. H., Leviton, A. & Dooling, E. C. The Developing Human Brain?: Growth and Epidemiologic Neuropathology. Elsevier Science, 1983).

29. Marconi, A. M. et al. Comparison of fetal and neonatal growth curves in detecting growth restriction. Obstet. Gynecol. 112, 1227–34 (2008).

30. Thomas, G. D. et al. Human Blood Monocyte Subsets. Arterioscler. Thromb. Vasc. Biol. 37, 1548–1558 (2017).

31. Ziegler-Heitbrock, L. et al. Nomenclature of monocytes and dendritic cells in blood. Blood 116, e74–e80 (2010).

32. Ertem, I. O. et al. A Guide for Monitoring Child Development in Low- and Middle-Income Countries. Pediatrics 121, e581–e589 (2008).

33. Ozturk Ertem, I. et al. Validation of the International Guide for Monitoring Child Development demonstrates good sensitivity and specificity in four diverse countries. Acta Paediatr. 108, 1074–1086 (2019).

34. Ali, S., Mustafa, S., Balaji, P., Dhaded, S. & Goudar, S. Guide for monitoring child development in Indian setting. IMRJ-Child Dev. 1, 05–07 (2011).

35. SAFOODS. FoodFinder 3.

36. Institute of Medicine (US) Committee to Review Dietary Reference Intakes for et al. Dietary Reference Intakes for Calcium and Vitamin D. Dietary Reference Intakes for Calcium and Vitamin D (National Academies Press (US), 2011). doi:10.17226/13050

37. Mchiza, Z. et al. A Review of Dietary Surveys in the Adult South African Population from 2000 to 2015. Nutrients 7, 8227–8250 (2015).

38. Kennedy, G., Ballard, T., Dop, M. C. (Marie C. & European Union. Guidelines for measuring household and individual dietary diversity. (Food and Agriculture Organization of the United Nations, 2011).

39. Labadarios, D., Steyn, N. P. & Nel, J. How diverse is the diet of adult South Africans? Nutr. J. 10, 33 (2011).

40. Ramokolo, V. et al. In Utero ART Exposure and Birth and Early Growth Outcomes Among HIV-Exposed Uninfected Infants Attending Immunization Services: Results From National PMTCT Surveillance, South Africa. Open Forum Infect. Dis. 4, ofx187 (2017).

41. Rosala-Hallas, A., Bartlett, J. W. & Filteau, S. Growth of HIV-exposed uninfected, compared with HIV-unexposed, Zambian children: a longitudinal analysis from infancy to school age. BMC Pediatr. 17, 80 (2017).

42. Evans, C., Chasekwa, B., Ntozini, R., Humphrey, J. H. & Prendergast, J. Head circumferences of children born to HIV-infected and HIV- uninfected mothers in Zimbabwe during the preantiretroviral therapy era. AIDS 30, 2323–2328 (2016).

43. Veena, S. R. et al. Association of birthweight and head circumference at birth to cognitive performance in 9-to 10-year-old children in South India: prospective birth cohort study. Pediatr. Res. 67, 424–9 (2010).

44. Eriksson, J. G., Kajantie, E., Lampl, M., Osmond, C. & Barker, D. J. P. Small head circumference at birth and early age at adiposity rebound. Acta Physiol. 210, 154–160 (2014).

45. Whitaker, R. C., Pepe, M. S., Wright, J. A., Seidel, K. D. & Dietz, W. H. Early adiposity rebound and the risk of adult obesity. Pediatrics 101, E5 (1998).

46. Risnes, K. R., Nilsen, T. I. L., Romundstad, P. R. & Vatten, L. J. Head size at birth and long-term mortality from coronary heart disease. Int. J. Epidemiol. 38, 955–962 (2009).

47. Hultman, C. M., Ohman, A., Cnattingius, S., Wieselgren, I. M. & Lindström, L. H. Prenatal and neonatal risk factors for schizophrenia. Br. J. Psychiatry 170, 128–33 (1997).

48. Williams, D. W. et al. CCR2 on CD14 ^+^ CD16 ^+^ monocytes is a biomarker of HIV-associated neurocognitive disorders. Neurol. - Neuroimmunol. Neuroinflammation 1, e36 (2014).

49. Conant, K. et al. Induction of monocyte chemoattractant protein-1 in HIV-1 Tat-stimulated astrocytes and elevation in AIDS dementia. Proc. Natl. Acad. Sci. U. S. A. 95, 3117–21 (1998).

50. Kelder, W., McArthur, J. C., Nance-Sproson, T., McClernon, D. & Griffin, D. E. ?-Chemokines MCP-1 and RANTES are selectively increased in cerebrospinal fluid of patients with human immunodeficiency virus-associated dementia. Ann. Neurol. 44, 831–835 (1998).

51. Prendergast, A. J. & Humphrey, J. H. The stunting syndrome in developing countries. Paediatr. Int. Child Health 34, 250–265 (2014).

52. Fadilah, A., Musson, R., Ong, M. T., Desurkar, A. V. & Mordekar, S. R. Vitamin B12 deficiency in infants secondary to maternal deficiency: A case series of seven infants. Eur. J. Paediatr. Neurol. EJPN 21, e3 (2017).

53. Webb-Girard, A. et al. Food insecurity is associated with attitudes towards exclusive breastfeeding among women in urban Kenya. Matern. Child Nutr. 8, 199–214 (2012).

54. Jama, N. A. et al. Enablers and barriers to success among mothers planning to exclusively breastfeed for six months: a qualitative prospective cohort study in KwaZulu-Natal, South Africa. Int. Breastfeed. J. 12, 43 (2017).

55. Lee, S. & Kelleher, S. L. Biological underpinnings of breastfeeding challenges: the role of genetics, diet, and environment on lactation physiology. Am. J. Physiol. Endocrinol. Metab. 311, E405–22 (2016).

56. WHO | Breast is always best, even for HIV-positive mothers. WHO (2011).

